# Inpatient Burden of Respiratory Syncytial Virus Infection and Influenza in Children Younger than 5 years in Japan, 2011-2022: A database study

**DOI:** 10.1101/2024.08.17.24312160

**Authors:** Takeshi Arashiro, Rolf Kramer, Jing Jin, Munehide Kano, Fangyuan Wang, Isao Miyairi

**Affiliations:** Sanofi Vaccines Medical, Tokyo, Japan; Faculty of Infectious and Tropical Diseases, London School of Hygiene and Tropical Medicine, London, United Kingdom; School of Tropical Medicine and Global Health, Nagasaki University, Nagasaki, Japan; Sanofi Vaccines Medical, Lyon, France; Sanofi Vaccines Medical, Beijing, China; Real World Evidence, Syneos Health, Beijing, China; Department of Pediatrics, School of Medicine, Hamamatsu University, Hamamatsu, Japan

**Keywords:** pediatrics, respiratory syncytial virus infections, influenza, hospitalization, Japan

## Abstract

**Background:** Respiratory syncytial virus (RSV) and influenza virus are major viral etiologies of pediatric lower respiratory tract infection, but comparative data on inpatient burden are lacking.

**Methods:** Using a large-scale health claims database in Japan, we identified patients under 5 years of age with a confirmed RSV or influenza diagnosis as an outpatient or inpatient between 2011-2022. Hospitalization rate, inpatient characteristics, various in-hospital outcomes/complications, and healthcare resource utilization were described.

**Results:** A total of 176,911 RSV-confirmed outpatients, 153,383 influenza-confirmed outpatients, 90,413 RSV-confirmed hospitalizations, and 11,186 influenza-confirmed hospitalizations were identified. Among outpatients, 24.7% of RSV infection and 2.8% of influenza cases required hospitalization within one week. There was no comorbidities/prematurity for 95.0% of RSV hospitalizations and 96.5% of influenza hospitalizations. Proportions of in-hospital outcomes/complications were (RSV infection vs influenza): oxygen use 47.6% vs 14.8%, mechanical ventilation 2.1% vs 0.7%, pneumonia 33.6% vs 12.8%, otitis media 7.7% vs 2.3%, febrile seizure 1.5% vs 34.4%, encephalitis/encephalopathy 0.1% vs 0.5%, myocarditis <0.1% vs 0.6%, antibiotics prescription 48.0% vs 24.4%. The mean inpatient stay was 6.1 vs 4.3 days at direct medical costs of 435,744 vs 315,809 JPY/patient. These trends held true in age-stratified data. In-hospital death occurred in 31 RSV infection and 6 influenza cases.

**Conclusions:** Although both infections resulted in substantial burden, RSV infection led to more frequent hospitalizations, worse in-hospital outcomes, longer inpatient stays, higher medical costs, and more frequent antibiotics prescription compared to influenza. Most RSV hospitalizations occurred among healthy term children, emphasizing the need for prevention measures in all children.

## Introduction

Lower respiratory tract infections (LRTIs) are major public health threats among young children. Among pediatric LRTIs, respiratory syncytial virus (RSV) and influenza virus are major viral etiologies with high burden globally.^1,2^ In Japan, the National Epidemiological Surveillance of Infectious Diseases (NESID) is conducted by the National Institute of Infectious Diseases (NIID) and Ministry of Health, Labour and Welfare (MHLW).^3^ For RSV infection and influenza, under the NESID, sentinel surveillance provides information on trends and levels of epidemics to inform policy, risk communication, and (for RSV infection) timing to initiate prophylaxis to individuals with underlying health conditions or prematurity.^4,5^ However, the NESID data only provides age-stratified aggregate data regardless of clinical severity. Therefore, detailed and comprehensive burden data on severe cases require special studies and such data are scarce in Japan. Most previous reports on severe cases of LRTI are based on small studies that describe single-center/single-prefecture experience or studies with only a brief description of in-hospital outcomess.^6–8^ Furthermore, although RSV and influenza share several clinical features in young children, comparative data on inpatient characteristics, inpatient outcomes, complications, and healthcare resource utilization (HCRU) is lacking globally. Finally, epidemiology of RSV infection and influenza has been disrupted globally due to the coronavirus disease (COVID-19) pandemic and further data especially around epidemic shift in age group are warranted.^9,10^ Therefore, we used one of the largest health claims databases in Japan to comprehensively describe RSV infection and influenza in children with the aim to elucidate hospitalization rate among infected individuals, inpatient characteristics, inpatient outcomes, and HCRU.

## Methods

### Database and study population

This retrospective cohort study was conducted using data from the Medical Data Vision (MDV) database, one of the largest electronic health records databases in Japan covering approximately 450 hospitals (25% of acute care hospitals in Japan). The present study population was comprised of patients under five years of age with a confirmed index diagnosis of RSV or influenza either outpatient or inpatient based on the International Classification of Diseases 10^th^ Revision (ICD-10) diagnostic codes (**Supplementary Table 1**) within an identification period of April 1, 2011 to July 31, 2022. Diagnoses which occurred within 6-months of the index diagnosis of the same infection (wash-out period) were excluded to avoid duplicating the same episode. Patients with a record of another infection with different pathogen within 30 days after the index date (e.g., suspected co-infection) were also excluded.

### Baseline characteristics

Data on age at diagnosis, sex, diagnosis date, and hospital size were collected. A 12-month look-back period from the index date was used to define the baseline period during which presence of comorbidities/prematurity and palivizumab use were ascertained. Comorbidities included in this study were those eligible to receive palivizumab in Japan (ICD-10 codes in **Supplementary Table 1**).^11–14^

### Definitions of hospitalized patients, clinical outcomes, healthcare resource utilization, and hospitalization rates

To describe inpatient characteristics including baseline characteristics, inpatient outcomes, and HCRU, we included all patients who had a confirmed diagnosis in the inpatient setting regardless of where or whether an outpatient RSV diagnosis was made (to ensure inclusion of patients who were diagnosed with RSV infection at different healthcare facilities such as clinics and patients who were never diagnosed before being hospitalized and diagnosed after hospitalization). To be identified as a relevant hospitalization, the infection had to have been coded as any of the following: (1) the disease that required the most medical resources, (2) primary disease for admission, or (3) disease that triggered the hospitalization. Patients who were admitted to the hospital were followed to examine in-hospital clinical outcomes and HCRU beginning from the index date and ending at the time of in-hospital death, discharge, the last record available for the patient, or the end of the study period (August 31, 2022), whichever occurred first. Variables for examination included intravenous (IV) fluid use, oxygen use, high flow-nasal canula oxygen therapy use, use of mechanical ventilation, intensive care unit (ICU) admissions, in-hospital death, antibiotics use, in-hospital complications (pneumonia, otitis media, febrile seizure, encephalitis or encephalopathy, and myocarditis). Finally, length of inpatient stay and direct medical costs during hospitalization for each patient were calculated by including all procedures, clinical exams, imaging, and medications (in Japanese yen).

For the purpose of calculating hospitalization rate among outpatients, we divided the number of patients with a confirmed index outpatient diagnosis by the number of patients who had a confirmed inpatient diagnosis in the same hospital. This was done to ensure that patients included in the numerator (hospitalized cases) were also in the denominator (outpatient cases). The basis of this algorithm was the assumption that almost all patients diagnosed in an outpatient clinic of a hospital will be admitted to the same hospital if the patient’s condition deteriorates to the point of requiring inpatient care, as per clinical practice in Japan. The date of first admission was used to estimate the percentage of eligible patients hospitalized within 0-6 days, 0-13 days, and 0-29 days of the index date. Percent hospitalizations within these time intervals were also calculated specifically for patients who required oxygen or intravenous (IV) fluids to restrict to those requiring medical attention.

### Analyses

We took a descriptive approach to understand baseline characteristics, hospitalizations rates, in-hospital clinical outcomes, and HCRU. We concluded that it would not be appropriate to estimate the relative measures of association (e.g., odds ratios) to compare such parameters between RSV infection and influenza, as various confounders (e.g., testing intensity) may exist. Instead, whenever possible, we presented data for the overall population, as well as data stratified by age since age is an important cofounder that influences various outcomes. Age categories used were less than 6 months, 6 months-1 year, 1-2 years, and 2-4 years of age (summary data were also presented by month for less than 1 year). Continuous variables were summarized with means/standard deviations (SD) and median/interquartile range (IQR). Categorical variables were summarized using counts and percentages.

Epidemic curves for outpatients and inpatients for RSV infection and influenza were created to describe temporal trends. Finally, to understand the influence of the COVID-19 pandemic on epidemiology, analyses were also performed and stratified by time period, including 2017-2019 (pre-COVID-19), 2020-2022 (during-COVID-19), and the 2021 season (as this season showed a unique RSV epidemic curve^3^). All analyses were performed using SAS software (version 9.4).

## Results

During the study period between April 2011 and July 2022, 176,911 RSV-confirmed cases and 153,383 influenza-confirmed cases were identified in the outpatient setting (**Figure 1; Supplementary Tables 2-3**). Furthermore, 90,413 RSV-confirmed hospitalizations and 11,186 influenza-confirmed hospitalizations were identified (overall **Table 1**; healthy children **Supplementary Table 4**). For both RSV infection and influenza, there were more males than females. Among hospitalized RSV cases, 39.0% were less than 6 months, 20.8% were 6 months-1 year, 26.4% were 1-2 years, and 13.7% were 2-4 years. Among hospitalized influenza cases, 19.1% were less than 6 months, 10.6% were 6 months-1 year, 25.4% were 1-2 years, and 44.8% were 2-4 years. Among inpatients, 85,896/90,413 (95.0%) of RSV cases and 10,789/11,186 (96.5%) of influenza cases occurred in patients with no record of relevant comorbidities or prematurity. **Supplementary Table 5** shows the data among infants by month of age.

**Figure 1.**
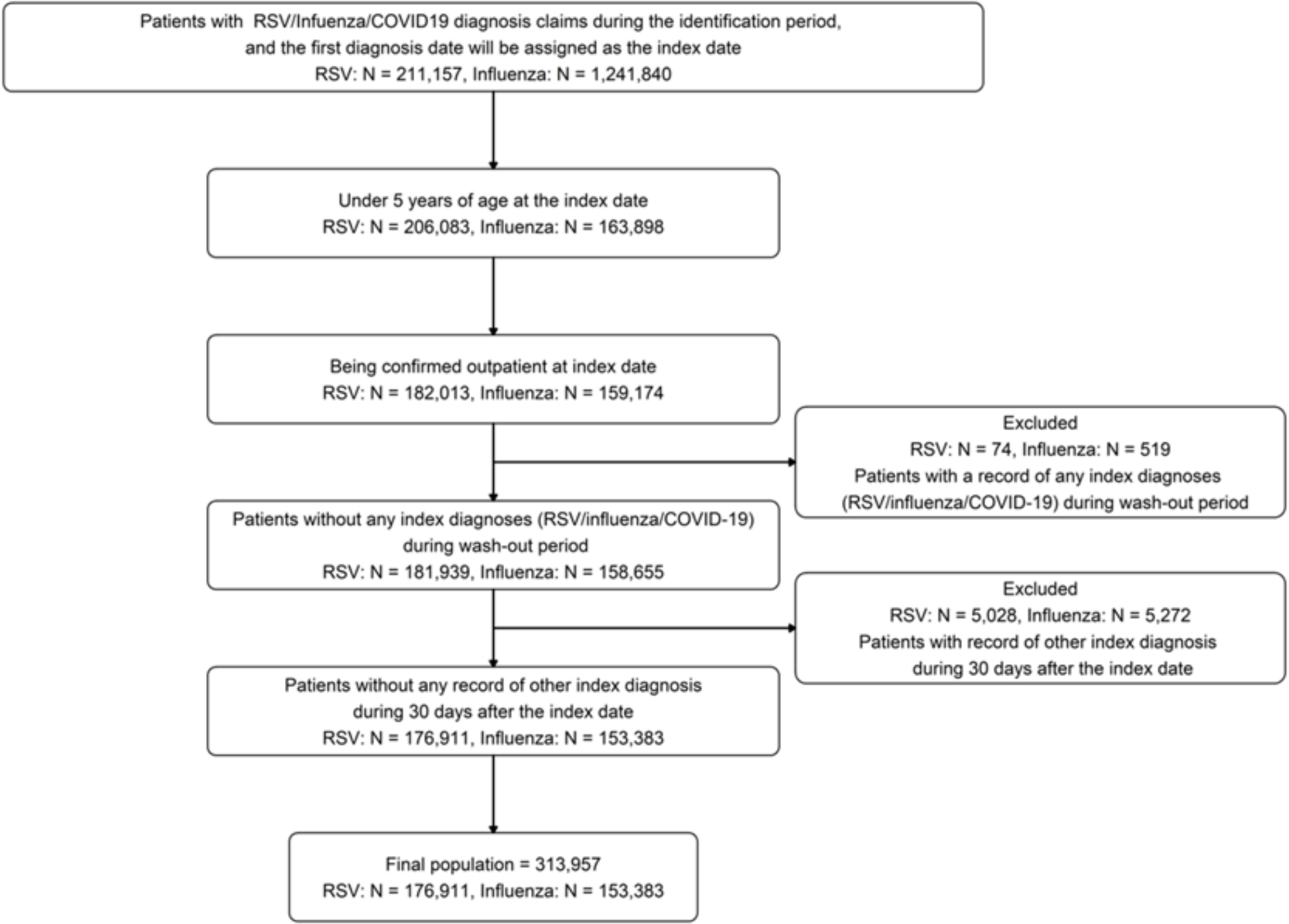
Description of the flow for assembling the outpatient population used to calculate hospital admission rates.

**Table 1.** Baseline characteristics for all children hospitalized with RSV infection and influenza overall and by age categories.

|  | RSV |  |  |  |  | Influenza |  |  |  |  |
| --- | --- | --- | --- | --- | --- | --- | --- | --- | --- | --- |
|  | Total | Age <6 months | Age 6-11 months | Age 1<2 years | Age 2<5 years | Total | Age <6 months | Age 6-11 months | Age 1<2 years | Age 2<5 years |
| **Total number of patients, n (relative % by age group)** | 90,413<br>(100) | 35,273<br>(39.0) | 18,827<br>(20.8) | 23,898<br>(26.4) | 12,415<br>(13.7) | 11,186<br>(100) | 2,141<br>(19.1) | 1,188<br>(10.6) | 2,845<br>(25.4) | 5,012<br>(44.8) |
| **Sex, n (%)** |  |  |  |  |  |  |  |  |  |  |
| Female | 40,958<br>(45.3) | 15,564<br>(44.1) | 8,030<br>(42.7) | 11,133<br>(46.6) | 6,231<br>(50.2) | 4,890<br>(43.7) | 986<br>(46.1) | 517<br>(43.5) | 1,282<br>(45.1) | 2,105<br>(42.0) |
| Male | 49,455<br>(54.7) | 19,709<br>(55.9) | 10,797<br>(57.4) | 12,765<br>(53.4) | 6,184<br>(49.8) | 6,296<br>(56.3) | 1,155<br>(54.0) | 671<br>(56.5) | 1,563<br>(54.9) | 2,907<br>(58.0) |
| **Age, months** |  |  |  |  |  |  |  |  |  |  |
| **Mean (SD)** | (11.8, 11.1) | (2.5, 1.4) | (8.4, 1.7) | (16.7, 3.4) | (33.9, 8.7) | (24.0, 17.4) | (2.6, 1.3) | (8.6, 1.7) | (17.4, 3.3) | (40.7, 10.6) |
| **Median (IQR)** | 9.0<br>(3.0-17.0) | 2.0<br>(1.0-4.0) | 8.0<br>(7.0-10.0) | 16.0<br>(14.0-19.0) | 32.0<br>(27.0-40.0) | 21.0<br>(9.0-38.0) | 2.0<br>(2.0-3.0) | 9.0<br>(7.0-10.0) | 17.0<br>(14.0-20.0) | 40.0<br>(31.0-50.0) |
| **Season by year, n (%)** |  |  |  |  |  |  |  |  |  |  |
| 2011 (May 2011 to April 2012) | 2,000<br>(2.2) | 724<br>(2.1) | 451<br>(2.4) | 547<br>(2.3) | 278<br>(2.2) | 360<br>(3.2) | 70<br>(3.3) | 38<br>(3.2) | 79<br>(2.8) | 173<br>(3.5) |
| 2012 (May 2012 to April 2013) | 3,609<br>(4.0) | 1,367<br>(3.9) | 845<br>(4.5) | 940<br>(3.9) | 457<br>(3.7) | 517<br>(4.6) | 99<br>(4.6) | 44<br>(3.7) | 129<br>(4.5) | 245<br>(4.9) |
| 2013 (May 2013 to April 2014) | 5,259<br>(5.8) | 2,252<br>(6.4) | 1,191<br>(6.3) | 1,227<br>(5.1) | 589<br>(4.8) | 933<br>(8.4) | 147<br>(6.9) | 99<br>(8.4) | 223<br>(7.9) | 464<br>(9.3) |
| 2014 (May 2014 to April 2015) | 8,057<br>(8.9) | 3,223<br>(9.1) | 1,793<br>(9.5) | 2,038<br>(8.5) | 1,003<br>(8.1) | 1,142<br>(10.2) | 262<br>(12.3) | 117<br>(9.9) | 275<br>(9.7) | 488<br>(9.8) |
| 2015 (May 2015 to April 2016) | 9,532<br>(10.6) | 3,834<br>(10.9) | 2,057<br>(10.9) | 2,464<br>(10.3) | 1,177<br>(9.5) | 1,575<br>(14.1) | 231<br>(10.8) | 202<br>(17.1) | 403<br>(14.2) | 739<br>(14.8) |
| 2016 (May 2016 to April 2017) | 8,926<br>(9.9) | 3,626<br>(10.3) | 2,057<br>(10.9) | 2,302<br>(9.6) | 941<br>(7.6) | 1,378<br>(12.4) | 302<br>(14.1) | 128<br>(10.8) | 348<br>(12.3) | 600<br>(12.0) |
| 2017 (May 2017 to April 2018) | 13,434<br>(14.9) | 5,148<br>(14.6) | 2,987<br>(15.9) | 3,588<br>(15.0) | 1,711<br>(13.8) | 1,677<br>(15.0) | 303<br>(14.2) | 174<br>(14.7) | 396<br>(13.9) | 804<br>(16.1) |
| 2018 (May 2018 to April 2019) | 12,356<br>(13.7) | 5,105<br>(14.5) | 2,754<br>(14.6) | 3,148<br>(13.2) | 1,349<br>(10.9) | 1,895<br>(17.0) | 423<br>(19.8) | 186<br>(15.7) | 490<br>(17.3) | 796<br>(16.0) |
| 2019 (May 2019 to April 2020) | 11,576<br>(12.8) | 4,380<br>(12.4) | 2,382<br>(12.7) | 3,313<br>(13.9) | 1,501<br>(12.1) | 1,660<br>(14.9) | 296<br>(13.9) | 194<br>(16.4) | 491<br>(17.3) | 679<br>(13.6) |
| 2020 (May to December) | 802<br>(0.9) | 304<br>(0.9) | 166<br>(0.9) | 238<br>(1.0) | 94<br>(0.8) | 3<br>(0.0) | 1<br>(0.1) | 2<br>(0.2) | 0<br>(0.0) | 0<br>(0.0) |
| 2021 (January to December) | 12,396<br>(13.7) | 4,456<br>(12.6) | 1,771<br>(9.4) | 3,438<br>(14.4) | 2,731<br>(22.0) | 6<br>(0.1) | 1<br>(0.1) | 1<br>(0.1) | 3<br>(0.1) | 1<br>(0.0) |
| 2022 (January to July) | 2,410<br>(2.7) | 832<br>(2.4) | 362<br>(1.9) | 638<br>(2.7) | 578<br>(4.7) | 4<br>(0.0) | 1<br>(0.1) | 0<br>(0.0) | 3<br>(0.1) | 0<br>(0.0) |
| **Comorbidity during baseline period, n (%)** |  |  |  |  |  |  |  |  |  |  |
| Down syndrome | 387 | 123 | 91 | 74 | 99 | 20 | 3 | 4 | 2 | 11 |
|  | (0.4) | (0.4) | (0.5) | (0.3) | (0.8) | (0.2) | (0.1) | (0.3) | (0.1) | (0.2) |
| Congenital heart disease | 1,817 | 902 | 501 | 264 | 150 | 119 | 36 | 33 | 29 | 21 |
|  | (2.0) | (2.6) | (2.7) | (1.1) | (1.2) | (1.1) | (1.7) | (2.8) | (1.0) | (0.4) |
| Bronchopulmonary dysplasia | 110 | 40 | 42 | 18 | 10 | 5 | 1 | 2 | 1 | 1 |
|  | (0.1) | (0.1) | (0.2) | (0.1) | (0.1) | (0.0) | (0.1) | (0.2) | (0.0) | (0.0) |
| Prematurity | 1,979 | 1,288 | 546 | 122 | 23 | 79 | 35 | 36 | 6 | 2 |
|  | (2.2) | (3.7) | (2.9) | (0.5) | (0.2) | (0.7) | (1.6) | (3.0) | (0.2) | (0.0) |
| Immunodeficiency | 169 | 68 | 45 | 35 | 21 | 29 | 4 | 6 | 5 | 14 |
|  | (0.2) | (0.2) | (0.2) | (0.2) | (0.2) | (0.3) | (0.2) | (0.5) | (0.2) | (0.3) |
| Pulmonary hypoplasia | 19 | 10 | 4 | 2 | 3 | 2 | 1 | 1 | 0 | 0 |
|  | (0.0) | (0.0) | (0.0) | (0.0) | (0.0) | (0.0) | (0.1) | (0.1) | (0.0) | (0.0) |
| Airway stenosis | 36 | 9 | 9 | 13 | 5 | 2 | 0 | 0 | 2 | 0 |
|  | (0.0) | (0.0) | (0.1) | (0.1) | (0.0) | (0.0) | (0.0) | (0.0) | (0.1) | (0.0) |
| Esophageal atresia | 3 | 1 | 0 | 2 | 0 | 0 | 0 | 0 | 0 | 0 |
|  | (0.0) | (0.0) | (0.0) | (0.0) | (0.0) | (0.0) | (0.0) | (0.0) | (0.0) | (0.0) |
| Congenital metabolic disorder | 378 | 82 | 100 | 115 | 81 | 93 | 3 | 9 | 32 | 49 |
|  | (0.4) | (0.2) | (0.5) | (0.5) | (0.7) | (0.8) | (0.1) | (0.8) | (1.1) | (1.0) |
| Neuromuscular disorder | 358 | 68 | 77 | 106 | 107 | 100 | 1 | 13 | 30 | 56 |
|  | (0.4) | (0.2) | (0.4) | (0.4) | (0.9) | (0.9) | (0.1) | (1.1) | (1.1) | (1.1) |
| Cystic fibrosis | 0 | 0 | 0 | 0 | 0 | 0 | 0 | 0 | 0 | 0 |
|  | (0.0) | (0.0) | (0.0) | (0.0) | (0.0) | (0.0) | (0.0) | (0.0) | (0.0) | (0.0) |
| No record of these comorbidities | 85,896 | 32,977 | 17,671 | 23,251 | 11,997 | 10,789 | 2,064 | 1,100 | 2,750 | 4,875 |
|  | (95.0) | (93.5) | (93.9) | (97.3) | (96.6) | (96.5) | (96.4) | (92.6) | (96.7) | (97.3) |
| **Palivizumab use, n (%)** |  |  |  |  |  |  |  |  |  |  |
| Yes | 1,559 | 706 | 453 | 354 | 46 | 82 | 16 | 28 | 33 | 5 |
|  | (1.7) | (2.0) | (2.4) | (1.5) | (0.4) | (0.7) | (0.8) | (2.4) | (1.2) | (0.1) |
| No record of use | 88,854 | 34,567 | 18,374 | 23,544 | 12,369 | 11,104 | 2,125 | 1,160 | 2,812 | 5,007 |
|  | (98.3) | (98.0) | (97.6) | (98.5) | (99.6) | (99.3) | (99.3) | (97.6) | (98.8) | (99.9) |
| **Size of healthcare facilities, n (%)** |  |  |  |  |  |  |  |  |  |  |
| Less than 200 beds | 5,252 | 1,781 | 1,209 | 1,454 | 808 | 551 | 68 | 55 | 156 | 272 |
|  | (5.8) | (5.1) | (6.4) | (6.1) | (6.5) | (4.9) | (3.2) | (4.6) | (5.5) | (5.4) |
| 200 to 499 beds | 50,876 | 19,056 | 11,029 | 13,859 | 6,932 | 6,090 | 1,138 | 640 | 1,578 | 2,734 |
|  | (56.3) | (54.0) | (58.6) | (58.0) | (55.8) | (54.4) | (53.2) | (53.9) | (55.5) | (54.6) |
| 500 or more beds | 34,285 | 14,436 | 6,589 | 8,585 | 4,675 | 4,545 | 935 | 493 | 1,111 | 2,006 |
|  | (37.9) | (40.9) | (35.0) | (35.9) | (37.7) | (40.6) | (43.7) | (41.5) | (39.1) | (40.0) |

*Rounding may create totals slightly over or below 100% in some cases.*

Regarding the epidemic curves for influenza, a clear seasonal pattern in wintertime was observed with no epidemic occurring in late 2020-2022 (**Figure 2; Supplementary Table 6**). In contrast, RSV infection also had epidemic peaks, but with considerable year-to-year variability. Specifically, before and during 2016, the peaks were from fall to winter. In 2017-2019, peaks began and ended earlier spanning summer to fall. There was no epidemic in 2020, but the largest wave was observed in 2021 peaking in early summer. These findings were in line with the corresponding NESID data. Also, importantly, trends and levels of epidemics were similar between inpatients and outpatients for both RSV infection and influenza, respectively.

**Figure 2.**
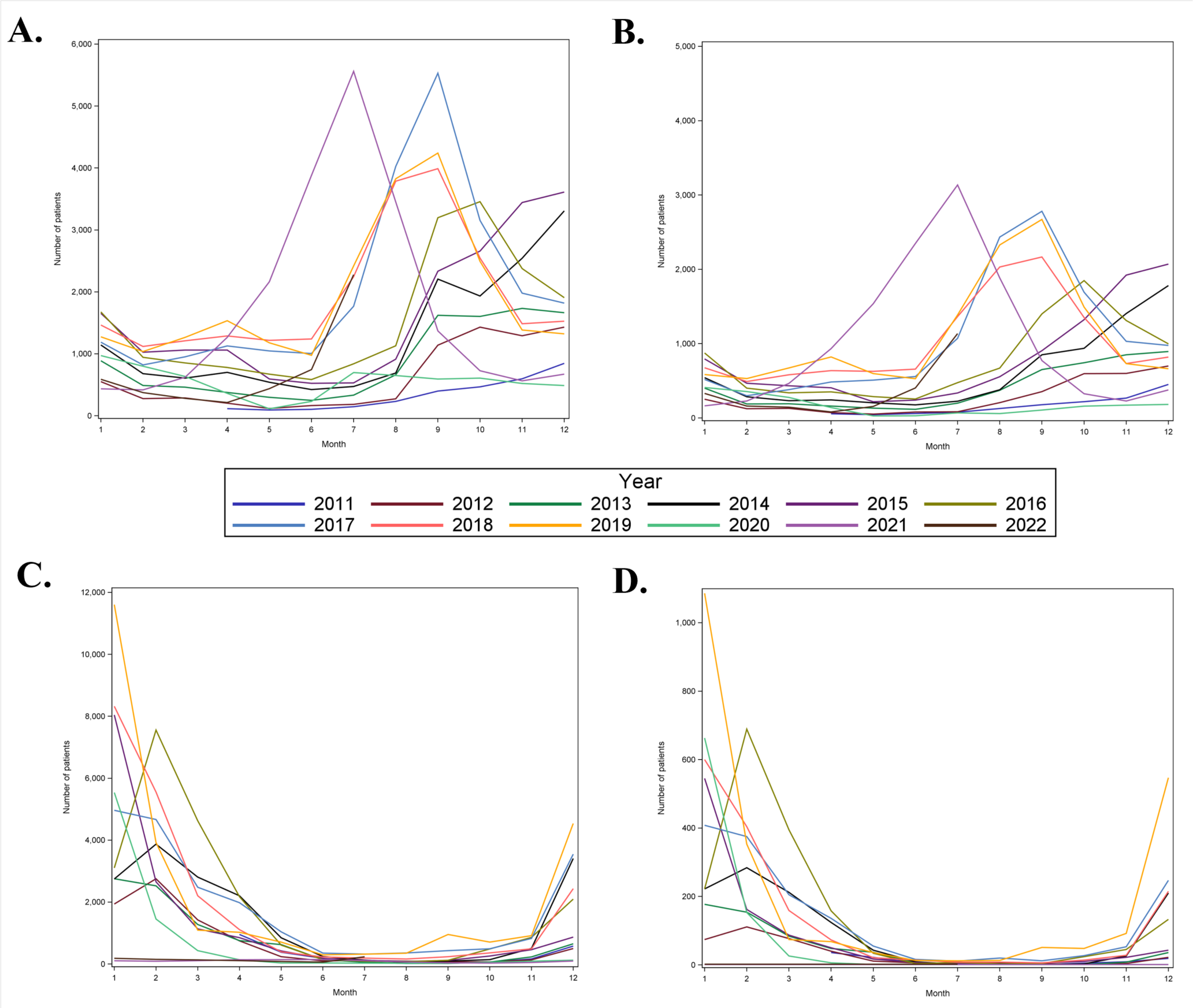
Epidemic curve for RSV and influenza identified in outpatient and inpatient settings. (A) Outpatient RSV infection, (B) inpatient RSV infection, (C) outpatient influenza, (D) inpatient influenza

Hospitalization rate among confirmed outpatient RSV diagnosis was 43,637/176,911 (24.7%) within 6 days of diagnosis (**Table 2**). This rate was similar when restricting to hospitalization requiring oxygen/IV usage (42,277/176,911; 23.9%). When stratified by age group, the highest percentage of hospitalization was observed among the 1-2 years age category (11,995/42,485; 28.2%), although this could be due to less outpatient testing (**Supplementary Methods**). Also, hospitalization rate within 6 days of diagnosis among healthy children was higher (41,666/141,843; 29.4%) than the non-restricted population and the highest percentage was observed for the less than 6 months age category within 29 days of diagnosis (15,717/45,318; 34.7%). In contrast, hospitalization rate among confirmed outpatient influenza diagnosis was much lower at 4,247/153,383 (2.8%) within 6 days of diagnosis. When stratified by age group, the highest percentage of hospitalization was observed among those less than 6 months age (736/8,741; 8.4%).

**Table 2.** Percentage of first hospitalizations after confirmed diagnosis of RSV or influenza at the outpatient setting in Japan among the overall population and after restricting to healthy children.

|  | RSV |  |  |  |  | Influenza |  |  |  |  |
| --- | --- | --- | --- | --- | --- | --- | --- | --- | --- | --- |
|  | Total | Age <6 months | Age 6-11 months | Age 1<2 years | Age 2<5 years | Total | Age <6 months | Age 6-11 months | Age 1<2 years | Age 2<5 years |
| **All Children** |  |  |  |  |  |  |  |  |  |  |
| **Total confirmed diagnoses at outpatient setting, n, (relative % by age group)** | 176,911<br>(100) | 70,021<br>(39.6) | 39,311<br>(22.2) | 42,485<br>(24.0) | 25,094<br>(14.2) | 153,383<br>(100) | 8,741<br>(5.7) | 16,295<br>(10.6) | 39,218<br>(25.6) | 89,129<br>(58.1) |
| **Overall hospitalizations after diagnosis (n, % of outpatient diagnoses)** |  |  |  |  |  |  |  |  |  |  |
| Admissions within 0-6 days | 43,637<br>(24.7) | 16,560<br>(23.7) | 8,903<br>(22.7) | 11,995<br>(28.2) | 6,179<br>(24.6) | 4,247<br>(2.8) | 736<br>(8.4) | 441<br>(2.7) | 1,126<br>(2.9) | 1,944<br>(2.2) |
| Admissions within 0-13 days | 43,805<br>(24.8) | 16,645<br>(23.8) | 8,949<br>(22.8) | 12,024<br>(28.3) | 6,187<br>(24.7) | 4,270<br>(2.8) | 739<br>(8.5) | 444<br>(2.7) | 1,135<br>(2.9) | 1,952<br>(2.2) |
| Admissions within 0-29 days | 44,281<br>(25.0) | 16,869<br>(24.1) | 9,090<br>(23.1) | 12,116<br>(28.5) | 6,206<br>(24.7) | 4,327<br>(2.8) | 740<br>(8.5) | 448<br>(2.8) | 1,156<br>(3.0) | 1,983<br>(2.2) |
| **Hospitalizations with oxygen/IV usage (n, % of outpatient diagnoses)** |  |  |  |  |  |  |  |  |  |  |
| Admissions within 0-6 days | 42,277<br>(23.9) | 15,713<br>(22.4) | 8,715<br>(22.2) | 11,841<br>(27.9) | 6,008<br>(23.9) | 4,089<br>(2.7) | 695<br>(8.0) | 426<br>(2.6) | 1,092<br>(2.8) | 1,876<br>(2.1) |
| Admissions within 0-13 days | 42,421<br>(24.0) | 15,787<br>(22.6) | 8,755<br>(22.3) | 11,865<br>(27.9) | 6,014<br>(24.0) | 4,112<br>(2.7) | 698<br>(8.0) | 429<br>(2.6) | 1,101<br>(2.8) | 1,884<br>(2.1) |
| Admissions within 0-29 days | 42,881<br>(24.2) | 15,998<br>(22.9) | 8,894<br>(22.6) | 11,956<br>(28.1) | 6,033<br>(24.0) | 4,167<br>(2.7) | 699<br>(8.0) | 433<br>(2.7) | 1,122<br>(2.9) | 1,913<br>(2.2) |
| **Healthy Children** |  |  |  |  |  |  |  |  |  |  |
| **Overall hospitalizations after diagnosis n, (relative % by age group)** | 141,843<br>(100) | 45,318<br>(31.9) | 33,225<br>(23.4) | 39,545<br>(27.9) | 23,755<br>(16.7) | 149,529<br>(100) | 8,197<br>(5.5) | 15,277<br>(10.2) | 38,019<br>(25.4) | 88,036<br>(58.9) |
| **Overall hospitalizations after diagnosis, n, (% of outpatient diagnoses)** |  |  |  |  |  |  |  |  |  |  |
| Admissions within 0-6 days | 41,666<br>(29.4) | 15,717<br>(34.7) | 8,443<br>(25.4) | 11,573<br>(29.3) | 5,933<br>(25.0) | 4,020<br>(2.7) | 694<br>(8.5) | 386<br>(2.5) | 1,066<br>(2.8) | 1,874<br>(2.1) |
| Admissions within 0-13 days | 41,814<br>(29.5) | 15,794<br>(34.9) | 8,479<br>(25.5) | 11,600<br>(29.3) | 5,941<br>(25.0) | 4,042<br>(2.7) | 697<br>(8.5) | 389<br>(2.5) | 1,075<br>(2.8) | 1,881<br>(2.1) |
| Admissions within 0-29 days | 42,228<br>(29.8) | 15,970<br>(35.2) | 8,610<br>(25.9) | 11,688<br>(29.6) | 5,960<br>(25.1) | 4,095<br>(2.7) | 698<br>(8.5) | 392<br>(2.6) | 1,094<br>(2.9) | 1,911<br>(2.2) |
| **Hospitalizations with oxygen/IV usage**<br>**(% of outpatient diagnoses)** |  |  |  |  |  |  |  |  |  |  |
| Admissions within 0-6 days | 40,384<br>(28.5) | 14,919<br>(32.9) | 8,276<br>(24.9) | 11,425<br>(28.9) | 5,764<br>(24.3) | 3,874<br>(2.6) | 657<br>(8.0) | 375<br>(2.5) | 1,033<br>(2.7) | 1,809<br>(2.1) |
| Admissions within 0-13 days | 40,515<br>(28.6) | 14,988<br>(33.1) | 8,308<br>(25.0) | 11,449<br>(29.0) | 5,770<br>(24.3) | 3,896<br>(2.6) | 660<br>(8.1) | 378<br>(2.5) | 1,042<br>(2.7) | 1,816<br>(2.1) |
| Admissions within 0-29 days | 40,916<br>(28.8) | 15,154<br>(33.4) | 8,437<br>(25.4) | 11,536<br>(29.2) | 5,789<br>(24.4) | 3,947<br>(2.6) | 661<br>(8.1) | 381<br>(2.5) | 1,061<br>(2.8) | 1,844<br>(2.1) |

*\*Healthy children were defined as those with children without a record of comorbidity of interest and no palivizumab use.*

Various in-hospital outcomes/HCRU-related factors were described (**Table 3**). IV fluid was used in 86,258/90,413 (95.4%) of RSV cases and 10,622/11,186 (95.0%) of influenza cases. Oxygen was required in 43,056/90,413 (47.6%) of RSV cases and 1,656/11,186 (14.8%) of influenza cases.

**Table 3.** Clinical outcomes and healthcare resource utilization and cost for children hospitalized with RSV infection and influenza.

|  | RSV |  |  |  |  | Influenza |  |  |  |  |
| --- | --- | --- | --- | --- | --- | --- | --- | --- | --- | --- |
|  | Total | Age <6 months | Age 6-11 months | Age 1<2 years | Age 2<5 years | Total | Age <6 months | Age 6-11 months | Age 1<2 years | Age 2<5 years |
| **Total number of patients, n (relative % by age group)** | 90,413<br>(100) | 35,273<br>(39.0) | 18,827<br>(20.8) | 23,898<br>(26.4) | 12,415<br>(13.7) | 11,186<br>(100) | 2,141<br>(19.1) | 1,188<br>(10.6) | 2,845<br>(25.4) | 5,012<br>(44.8) |
| **Intravenous fluids use** |  |  |  |  |  |  |  |  |  |  |
| No | 4,155<br>(4.6) | 2,432<br>(6.9) | 656<br>(3.5) | 548<br>(2.3) | 519<br>(4.2) | 564<br>(5.0) | 124<br>(5.8) | 58<br>(4.9) | 122<br>(4.3) | 260<br>(5.2) |
| Yes | 86,258<br>(95.4) | 32,841<br>(93.1) | 18,171<br>(96.5) | 23,350<br>(97.7) | 11,896<br>(95.8) | 10,622<br>(95.0) | 2,017<br>(94.2) | 1,130<br>(95.1) | 2,723<br>(95.7) | 4,752<br>(94.8) |
| **Oxygen use** |  |  |  |  |  |  |  |  |  |  |
| No | 47,357<br>(52.4) | 19,575<br>(55.5) | 9,106<br>(48.4) | 12,105<br>(50.7) | 6,571<br>(52.9) | 9,530<br>(85.2) | 2,014<br>(94.1) | 1,016<br>(85.5) | 2,292<br>(80.6) | 4,208<br>(84.0) |
| Yes | 43,056<br>(47.6) | 15,698<br>(44.5) | 9,721<br>(51.6) | 11,793<br>(49.4) | 5,844<br>(47.1) | 1,656<br>(14.8) | 127<br>(5.9) | 172<br>(14.5) | 553<br>(19.4) | 804<br>(16.0) |
| **High flow nasal canula oxygen therapy use** |  |  |  |  |  |  |  |  |  |  |
| No | 88,117<br>(97.5) | 33,791<br>(95.8) | 18,426<br>(97.9) | 23,614<br>(98.8) | 12,286<br>(99.0) | 11,162<br>(99.8) | 2,137<br>(99.8) | 1,185<br>(99.8) | 2,841<br>(99.9) | 4,999<br>(99.7) |
| Yes | 2,296<br>(2.5) | 1,482<br>(4.2) | 401<br>(2.1) | 284<br>(1.2) | 129<br>(1.0) | 24<br>(0.2) | 4<br>(0.2) | 3<br>(0.3) | 4<br>(0.1) | 13<br>(0.3) |
| **Use of mechanical ventilation** |  |  |  |  |  |  |  |  |  |  |
| No | 88,485<br>(97.9) | 34,118<br>(96.7) | 18,510<br>(98.3) | 23,593<br>(98.7) | 12,264<br>(98.8) | 11,112<br>(99.3) | 2,133<br>(99.6) | 1,183<br>(99.6) | 2,826<br>(99.3) | 4,970<br>(99.2) |
| Yes | 1,928<br>(2.1) | 1,155<br>(3.3) | 317<br>(1.7) | 305<br>(1.3) | 151<br>(1.2) | 74<br>(0.7) | 8<br>(0.4) | 5<br>(0.4) | 19<br>(0.7) | 42<br>(0.8) |
| **Intensive care unit (ICU) admissions** |  |  |  |  |  |  |  |  |  |  |
| No | 89,813<br>(99.3) | 34,842<br>(98.8) | 18,743<br>(99.6) | 23,846<br>(99.8) | 12,382<br>(99.7) | 11,147<br>(99.7) | 2,138<br>(99.9) | 1,186<br>(99.8) | 2,831<br>(99.5) | 4,992<br>(99.6) |
| Yes | 600<br>(0.7) | 431<br>(1.2) | 84<br>(0.5) | 52<br>(0.2) | 33<br>(0.3) | 39<br>(0.4) | 3<br>(0.1) | 2<br>(0.2) | 14<br>(0.5) | 20<br>(0.4) |
| **In-hospital death** |  |  |  |  |  |  |  |  |  |  |
| No | 90,382<br>(100) | 35,262<br>(100) | 18,818<br>(100) | 23,892<br>(100) | 12,410<br>(100) | 11,180<br>(100) | 2,140<br>(100) | 1,188<br>(100) | 2,844<br>(100) | 5,008<br>(99.9) |
| Yes | 31<br>(0.0) | 11<br>(0.0) | 9<br>(0.1) | 6<br>(0.0) | 5<br>(0.0) | 6<br>(0.1) | 1<br>(0.1) | 0<br>(0.0) | 1<br>(0.0) | 4<br>(0.1) |
| **Antibiotics use** |  |  |  |  |  |  |  |  |  |  |
| No | 46,995<br>(52.0) | 23,461<br>(66.5) | 8,854<br>(47.0) | 9,337<br>(39.1) | 5,343<br>(43.0) | 8,458<br>(75.6) | 1,792<br>(83.7) | 866<br>(72.9) | 2,067<br>(72.7) | 3,733<br>(74.5) |
| Yes | 43,418<br>(48.0) | 11,812<br>(33.5) | 9,973<br>(53.0) | 14,561<br>(60.9) | 7,072<br>(57.0) | 2,728<br>(24.4) | 349<br>(16.3) | 322<br>(27.1) | 778<br>(27.4) | 1,279<br>(25.5) |
| **In-hospital complications** |  |  |  |  |  |  |  |  |  |  |
| Pneumonia | 30,376 | 6,590 | 6,484 | 11,353 | 5,949 | 1,428 | 115 | 158 | 413 | 742 |
|  | (33.6) | (18.7) | (34.4) | (47.5) | (47.9) | (12.8) | (5.4) | (13.3) | (14.5) | (14.8) |
| Otitis media | 6,915 | 1,047 | 1,876 | 2,911 | 1,081 | 255 | 17 | 45 | 102 | 91 |
|  | (7.7) | (3.0) | (10.0) | (12.2) | (8.7) | (2.3) | (0.8) | (3.8) | (3.6) | (1.8) |
| Febrile seizure | 1,366 | 67 | 130 | 761 | 408 | 3,843 | 44 | 254 | 1,430 | 2,115 |
|  | (1.5) | (0.2) | (0.7) | (3.2) | (3.3) | (34.4) | (2.1) | (21.4) | (50.3) | (42.2) |
| Encephalitis or encephalopathy | 49 | 24 | 2 | 12 | 11 | 54 | 4 | 5 | 24 | 21 |
|  | (0.1) | (0.1) | (0.0) | (0.1) | (0.1) | (0.5) | (0.2) | (0.4) | (0.8) | (0.4) |
| Myocarditis | 4 | 1 | 1 | 1 | 1 | 68 | 1 | 5 | 15 | 47 |
|  | (0.0) | (0.0) | (0.0) | (0.0) | (0.0) | (0.6) | (0.1) | (0.4) | (0.5) | (0.9) |
| **Length of inpatient stay, days** |  |  |  |  |  |  |  |  |  |  |
| **Mean (SD)** | 6.1 (3.3) | 6.3 (3.7) | 6.2 (2.9) | 5.9 (3.4) | 5.7 (2.5) | 4.3 (2.6) | 4.1 (1.8) | 4.5 (2.8) | 4.4 (2.5) | 4.3 (2.8) |
| **Median (IQR)** | 6.0<br>(4.0-7.0) | 6.0<br>(4.0-7.0) | 6.0<br>(4.0-7.0) | 5.0<br>(4.0-7.0) | 5.0<br>(4.0-7.0) | 4.0<br>(3.0-5.0) | 4.0<br>(3.0-4.0) | 4.0<br>(3.0-5.0) | 4.0<br>(3.0-5.0) | 4.0<br>(3.0-5.0) |
| **Total costs during hospitalization (per patient)** |  |  |  |  |  |  |  |  |  |  |
| **Mean (SD)** | 435,743.8<br>(271,703.1) | 453,731.5<br>(314,856.1) | 434,766.4<br>(248,195.8) | 420,035.9<br>(243,875.7) | 416,362.1<br>(215,192.6) | 315,809.4<br>(209,438.7) | 292,933.0<br>(151,420.1) | 327,031.6<br>(248,576.5) | 330,289.5<br>(234,427.2) | 314,702.2<br>(204,736.6) |
| **Median (IQR)** | 391,648.5<br>(286,581.3-524,253.1) | 402,215.5<br>(291,487.7-542,645.5) | 391,262.3<br>(286,157.9-524,239.2) | 3847,67.1<br>(283,405.0-506,331.2) | 380722.7<br>(280912.3-504248.2) | 2821,85.4<br>(207,122.7-371,700.4) | 272,527.0<br>(200,339.0-346,756.2) | 287,527.1<br>(217,363.1-377,209.5) | 295,797.2<br>(219,178.9-391,632.7) | 279,503.3<br>(202,282.5-372,237.4) |

All costs are noted in Japanese Yen, Rounding may create totals slightly over or below 100% in some cases; Abbreviations: IQR, interquartile range; SD, standard deviation

Mechanical ventilation was required in 1,928/90,413 (2.1%) of RSV cases and 74/11,186 (0.7%) of influenza cases. In-hospital death occurred in 31/90,413 (<0.1%) of RSV cases and 6/11,186 (0.1%) of influenza cases. Antibiotics were prescribed in 43,418/90,413 (48.0%) of RSV cases and 2,728/11,186 (24.4%) of influenza cases. The mean inpatient stays were 6.1 days and 4.3 days with direct medical costs of 435,744JPY/patient and 315,809JPY/patient for RSV infection and influenza, respectively. Regarding in-hospital complications, pneumonia occurred in 30,376/90,413 (33.6%) of RSV cases and 1,428/11,186 (12.8%) of influenza cases. Otitis media occurred in 6,915/90,413 (7.7%) of RSV cases and 255/11,186 (2.3%) of influenza cases. Febrile seizure occurred in 1,366/90,413 (1.5%) of RSV cases and 3,843/11,186 (34.4%) of influenza cases. Encephalitis or encephalopathy occurred in 49/90,413 (0.1%) of RSV cases and 54/11,186 (0.5%) of influenza cases. Myocarditis occurred in 4/90,413 (<0.1%) of RSV cases and 68/11,186 (0.6%) of influenza cases.

Finally, we stratified the number of hospitalizations by COVID-19 era (**Supplementary Tables 7-8**). There was a comparatively lower proportion of RSV hospitalizations in age <6 months and age 6-11 months during COVID-19 era (36.4%, 15.1%) and 2021 season (35.9%, 14.3%) compared to pre-COVID-19 era (39.3%, 21.9%). Conversely, there was a comparatively higher proportion of RSV hospitalizations in age 2 years to 4 years during COVID-19 era (21.3%) and 2021 season (22.0%) compared to pre-COVID-19 era (12.0%). There were very few influenza hospitalizations during COVID-19 era.

## Discussion

We conducted a study using large electronic health records database covering approximately 25% of acute care hospitals in Japan to elucidate the comprehensive inpatient burden of RSV infection and influenza in young children. We found that a large number of RSV and influenza cases were diagnosed in both outpatient and inpatient settings with substantial in-hospital outcomes/complications and HCRU during the 11-year study period.

Similar to what has been observed in previous reports^1^, over half of RSV hospitalizations occurred in infants less than 1 year of age. Approximately 95% of these RSV hospitalizations occurred in patients with no record of comorbidities or prematurity. We also observed hospitalization rates among children diagnosed with RSV infection to be 20-30%, meaning that one in four medically-attended RSV infections resulted in hospitalization. When restricting to patients without comorbidities/prematurity, the rates were even higher (up to 35%), which is likely to be due to unavailability of prevention measures in this population (unlike palivizumab for children with comorbidities/prematurity). These were in stark contrast to low hospitalization rates for influenza where only 2-3% in children 6 months of age or older and 8-9% among infants less 6 months of age.

We further examined in-hospital outcomes and HCRU related to each infection. Important in-hospital outcomes including oxygen use, ICU admission, and mechanical ventilation were more prevalent with RSV infections compared to influenza across all age groups. Still, ICU admission and mechanical ventilation among the RSV inpatient population were much lower compared to the data from the U.S., which could be due to a lower threshold of hospitalization in Japan and scarcity of pediatric ICUs in Japan (i.e., some specialized care including mechanical ventilation is routinely done in general wards).^15^ Although the numbers and proportions were small, a few in-hospital deaths occurred for both infections. Antibiotic use was more prevalent for RSV infection than for influenza. This could be due to more severe in-hospital outcomes for RSV infection and improved antimicrobial stewardship is warranted. Regarding in-hospital complications, pneumonia and otitis media were more common in RSV infection than in influenza. In contrast, febrile seizures, encephalitis/encephalopathy, and myocarditis were more common in influenza than in RSV infection. Notably, approximately one-third of influenza hospitalization were associated with febrile seizure. Longer inpatient stays (6.1 vs 4.3 days) with higher direct medical costs (435,744 vs 315,809 JPY/patient) were observed with RSV infection compared with influenza. Finally, analyses by COVID-19 era revealed a slight shift in age of RSV hospitalizations to older age groups in 2021-2022, which is in line with lack of epidemic in 2020. This shift may recover to pre-COVID-19 trends after a few years of epidemics together with disrupted seasonal pattern as observed in the U.S.^10^

Given the substantial burden of RSV infection with considerable in-hospital outcomes/complications and HCRU, the currently available RSV prophylaxis, palivizumab, which only covers children with specific comorbidities or prematurity, is not enough to protect the majority of children from hospitalization when infected, similar to influenza where routine vaccination is available. Novel RSV prevention measures to protect all children are needed. Recently, another monoclonal antibody with extended half-life, nirsevimab, has been approved for use as a preventative treatment for all infants entering their first RSV season in the U.S. and several other countries.^16^ Maternal immunization is also approved for use, although only available in a seasonal manner with a narrow window period for immunization (pregnant individuals at 32-36 weeks gestational age) in the U.S.^17^ There are other potential preventive measures for toddlers under development.^18^ The current study highlights that the burden of severe RSV infection in children in Japan and other countries are similarly. Therefore, improved awareness of disease burden as well as wide use of these preventive measures are highly anticipated upon approval.

### Limitations

There are several limitations to this study. First, the current database is based on participating hospitals only and there is no linkage of data across hospitals. Second, although both RSV and influenza testing is covered by NHI (**Supplementary Methods**), careful interpretation is necessary as testing intensity may differ between RSV infection and influenza as well as by age group (especially infants vs young children >1 years). Third, the database did not have information on influenza vaccination/anti-viral use, which may have influenced study outcomes. Nevertheless, the availability of prevention/treatment for influenza and unavailability to prevention/treatment for RSV infection is reflection of real-world disease burden from RSV disease, which may improve with novel prevention measures mentioned above. Finally, similar to other database studies, medical coding may not always reflect true diagnosis or procedures.

## Conclusions

A large number of RSV and influenza cases were diagnosed in both Japanese outpatient and inpatient settings during the study period. There were also substantial in-hospital outcomes/complications and HCRU, many of which were more frequently observed with RSV infection compared to influenza. Furthermore, longer inpatient stays with more expensive direct medical costs were seen with RSV infection compared to influenza. Approximately one in four RSV infection resulted in hospitalization with most hospitalization occurring among healthy term children, emphasizing the need for RSV prevention measures in all children.

## Data Availability

All data produced in the present work are contained in the manuscript.

## Ethics Statement

The study was conducted under the ethical principles consistent with the Japanese Ethical Guidelines for Medical and Biological Research Involving Human Subjects and the Declaration of Helsinki. The requirement for informed consent was waived as all personal and identifying data were anonymized in the administrative health claims dataset.

## Author Contributions

All authors contributed to the conception and/or design of the study; TA and FW were involved in data analyses; TA, RK, and IM were involved in interpretation of the data; TA drafted the first version of the manuscript. All authors critically reviewed the manuscript for intellectual content and gave final approval of the version to be published.

## Acknowledgements

We thank employees of Syneos Health for technical assistance in study design and data analysis. We also thank Ko Nakajo for technical assistance in study conception and design.

