## Supplementary Material for "Inpatient Burden of Respiratory Syncytial Virus Infection and Influenza in Children Younger than 5 years in Japan, 2011-2022: A database study"

**Supplementary Methods**

*RSV and influenza testing in Japan*

In Japan, RSV and influenza tests are covered by national health insurance (NHI) and most healthcare facilities use rapid antigen kit for testing. For RSV testing any of the following groups are covered under NHI: (1) outpatient infants less than 1 year of age, (2) any hospitalized child, or (3) children eligible for palivizumab (note that the test is covered by NHI if any one of these criteria applies). For influenza testing, any outpatient is covered by insurance, regardless of age.

*Influenza vaccination coverage and influenza antiviral use in Japan*

Influenza vaccination coverage among children in Japan is approximately 50-60%, which is similar to the United States (U.S.).[1,2] Outpatient antiviral use for influenza is reported to be more common among young children diagnosed for influenza in Japan (approximately 80%) compared to in the U.S. (approximately 40%).[3,4]

*Database*

This retrospective cohort study was conducted using data from the Medical Data Vision (MDV) database, one of the largest electronic health records databases in Japan covering approximately 450 hospitals (25% of acute care hospitals in Japan) which includes data for over 40 million people. The MDV database includes detailed data on inpatient and outpatient encounters, drugs prescribed, diagnoses, laboratory tests performed, and medical practice (e.g., date of visit, department visited) from these hospitals.

*Definition of seasonality*

Seasons were defined for each year based on the epidemic curves for RSV observed annually between 2011 and 2022 by the NESID. Seasons for 2011 to 2019 were defined as beginning on May 1 of each year and ending on April 30 of the following year. Due to COVID-19 pandemic, May to December for 2020 was defined as a non-epidemic period. Since RSV seasonality was irregular after 2021, 2021 and 2022 seasons were defined as January to December (note: the study period ended on July 31, 2022, so the 2022 season was only half as long as a full season).

*Definition of healthy infants*

Since the MDV database may over-represent individuals with comorbidities/prematurity given it is data from acute care hospitals, some analyses were also performed by restricting to a population of "healthy children" who had no claim of the select comorbidities or prematurity (described above) and no palivizumab use.

*Data on COVID-19*

We initially included analysis of COVID-19 as part of the study protocol. However, data on COVID-19 were excluded from the analyses as there were very few related hospitalizations (n=14) which may be because hospitalization with COVID-19 was relatively rare during the study period and these patients were concentrated in certain hospitals that had capacity to admit COVID-19 patients (e.g., pediatric COVID-19 ward). It is likely many of these hospitals were not included in the database.

*References*

1. Ministry of Health Labour and Welfare. About the Seasonal Influenza Vaccine for the 2023/2024 Season. https://www.mhlw.go.jp/content/10906000/001146456.pdf. Accessed December 23, 2023. (In Japanese)
2. Centers for Disease Control and Prevention. Flu Vaccination Coverage, United States, 2022–23 Influenza Season. https://www.cdc.gov/flu/fluvaxview/coverage-2223estimates.htm. Accessed December 23, 2023.
3. Okubo Y, Uda K, Miyairi I. Trends in Influenza and Related Health Resource Use During 2005–2021 Among Children in Japan. J Pediatr Infect Dis 2023;42:648-53.
4. Antoon JW, Sarker J, Abdelaziz A, et al. Trends in outpatient influenza antiviral use among children and adolescents in the United States. Pediatrics. 2023;152(6):e2023061960.

**Supplementary Table 1.** Relevant ICD-10 codes for conditions of interest

| Condition | ICD-10 codes | Detailed description |
|---|---|---|
| **Index disease** | | |
| RSV | J12.1 | Respiratory syncytial virus pneumonia |
| RSV | J20.5 | RSV bronchitis |
| RSV | J21.0 | RSV bronchiolitis |
| RSV | B34.8 | RSV infection |
| RSV | B97.4 | RSV bronchitis |
| Influenza | J09 | Novel influenza (H1N1) |
| Influenza | J10.1 | Influenza A/USSR |
| Influenza | J11.0 | Influenza pneumonia |
| Influenza | J11.1 | Influenza |
| **Comorbidities of interest** | | |
| Down syndrome | Q90.9 | |
| Asthma | J45.9 | |
| Bronchitis | J40 | |
| Congenital heart disease | I27.8, I51.0, Q20.0-5, Q20.8-Q21.4, Q21.8-Q22.6, Q22.8, Q23.0-4, Q23.8-Q24.4, Q24.5, Q24.8-9, Q25.1, Q25.3-Q264, Q26.8, Q26.9 | |
| Bronchopulmonary dysplasia | P27.1 | |
| Prematurity | P07.3 | |
| Immunodeficiency | D70.0, D71.0, D72.0, D84.9, D80.0-2, D80.4, D80.5, D80.9, D81.0, D81.9, D82.0, D82.1, D82.3, D83.0, D83.9, D84.0, D84.8, D89.1, D89.8, M35.9, Z21.0, Z94.0, Z94.1, Z94.2, Z94.3, Z94.4, Z94.8 | |
| Intellectual developmental delay | F70-F79 | |
| Children under supportive medical care | Japanese medical segment (kubun) codes: C101-2 – C116 | |
| Pulmonary hypoplasia | Q33.6 | |
| Airway stenosis | J98.8 | |
| Esophageal atresia | Q39.3 | |
| Congenital metabolic disorders | D52.8, E55.21, E70.0, E70.1, E70.2, E70.9, E71.0, E71.1, E71.3 E72.0, E72.1, E72.2, E72.3, E72.4, E72.9, E73.0, E74.0, E74.2, E74.4, E74.8, E74.9, E75.2, E75.5, E77.0, E77.8, E77.9, E78.0, E78.9, E79.0, E79.8, E83.0, E88.0, E88.8, E88.9, G60.1, G71.3, G90.8. P09.1 | |
| Cystic fibrosis | E84.9 | |
| Neuromuscular disorders | G10-14, G60, G70-73, G80-83, G90-99 | |
| **Complications** | | |
| Pneumonia | A40.3, A41.9, A49.1, G00.1, I30.1, J11.0, J12.1, J12.9, J13, J14, J15.1, J15.2, J15.4, J15.6, J15.9, J18.0, J18.1, J18.8, J18.9, J20.2, J69.0, J84.1, J84.9, J85.0, J85.1, J95.8, K65.0, U07.1 | |
| Otitis media | H65.0, H66.4, H66.0, H66.9, H65.1, H65.9, H67.0, H67.1 | |
| Febrile seizure | R56.0 | |
| Encephalitis or encephalopathy | G00-G09, G41 | |

| | |
|---|---|
| Myocarditis | J11.8, I40.0, I40.9, I51.4 |

**Note:** **ICD-10 coding limited to codes used by the Japanese healthcare system to identify the conditions of interest and are covered in the dataset*

**Supplementary Table 2.** Baseline characteristics of all children <5 years of age who were diagnosed in the outpatient setting with RSV infection and influenza

| | RSV | | | | | Influenza | | | | |
|---|---|---|---|---|---|---|---|---|---|---|
| | Total | Age <6 months | Age 6-11 months | Age 1<2 years | Age 2<5 years | Total | Age <6 months | Age 6-11 months | Age 1<2 years | Age 2<5 years |
| **Total number of patients, n relative % by age group)** | 176, 911 (100) | 70,021 (39.6) | 39,311 (22.2) | 42,485 (24.0) | 25,094 (14.2) | 153,383 (100) | 8,741 (5.7) | 16,295 (10.6) | 39,218 (25.6) | 89,129 (58.1) |
| **Sex, n (%)** | | | | | | | | | | |
| Female | 79,746 (45.1) | 30,989 (44.3) | 17,235 (43.8) | 19,506 (45.9) | 12,016 (47.9) | 70,589 (46.0) | 3,881 (44.4) | 7,476 (45.9) | 18,156 (46.3) | 41,076 (46.1) |
| Male | 97,165 (54.9) | 39,032 (55.7) | 22,076 (56.2) | 22,979 (54.1) | 13,078 (52.1) | 82,794 (54.0) | 4,860 (55.6) | 8,819 (54.1) | 21,062 (53.7) | 48,053 (53.9) |
| **Age, months** | | | | | | | | | | |
| **Mean (SD)** | 11.8 (11.6) | 2.5 (1.5) | 8.3 (1.7) | 16.6 (3.4) | 35.0 (9.2) | 29.8 (16.8) | 3.1 (1.4) | 8.8 (1.7) | 17.1 (3.4) | 41.9 (10.5) |
| **Median (IQR)** | 8.0 (3.0-17.0) | 2.0 (1.0-4.0) | 8.0 (7.0-10.0) | 16.0 (14.0-19.0) | 33.0 (27.0-41.0) | 28.0 (15.0-45.0) | 3.0 (2.0-4.0) | 9.0 (7.0-10.0) | 17.0 (14.0-20.0) | 42.0 (33.0-51.0) |
| **Season by year, n (%)** | | | | | | | | | | |
| 2011 (May 2011 to April 2012) | 4,191 (2.4) | 1,344 (1.9) | 877 (2.2) | 1,199 (2.8) | 771 (3.1) | 8,308 (5.5) | 198 (2.3) | 600 (3.7) | 1,816 (4.7) | 5,694 (6.4) |
| 2012 (May 2012 to April 2013) | 8,239 (4.7) | 2,985 (4.3) | 2,052 (5.2) | 2,022 (4.8) | 1,180 (4.7) | 8,619 (5.7) | 312 (3.6) | 721 (4.5) | 1,998 (5.1) | 5,588 (6.3) |
| 2013 (May 2013 to April 2014) | 11,285 (6.4) | 4,545 (6.5) | 2,756 (7.0) | 2,559 (6.0) | 1,425 (5.7) | 13,432 (8.8) | 523 (6.0) | 1,224 (7.6) | 3,334 (8.5) | 8,351 (9.4) |
| 2014 (May 2014 to April 2015) | 16,909 (9.6) | 6,678 (9.5) | 3,946 (10.0) | 3,958 (9.3) | 2,327 (9.3) | 18,084 (11.9) | 915 (10.5) | 1,754 (10.8) | 4,590 (11.8) | 10,825 (12.2) |
| 2015 (May 2015 to April 2016) | 18,853 (10.7) | 7,362 (10.5) | 4,596 (11.7) | 4,433 (10.4) | 2,462 (9.8) | 19,960 (13.1) | 858 (9.8) | 2,061 (12.7) | 5,055 (13.0) | 11,986 (13.6) |
| 2016 (May 2016 to April 2017) | 18,242 (10.3) | 7,330 (10.5) | 4,351 (11.1) | 4,318 (10.2) | 2,243 (9.0) | 18,560 (12.2) | 1,096 (12.6) | 2,038 (12.6) | 4,978 (12.8) | 10,448 (11.8) |

| | | | | | | | | | | |
|---|---|---|---|---|---|---|---|---|---|---|
| 2017 (May 2017 to April 2018) | 25,404 (14.4) | 9,486 (13.6) | 6,048 (15.4) | 6,346 (15.0) | 3,524 (14.1) | 24,544 (16.1) | 1,496 (17.2) | 2,785 (17.2) | 6,232 (16.0) | 14,031 (15.9) |
| 2018 (May 2018 to April 2019) | 23,143 (13.1) | 9,561 (13.7) | 5,315 (13.5) | 5,499 (13.0) | 2,768 (11.0) | 22,129 (14.5) | 1,683 (19.3) | 2,575 (15.9) | 5,706 (14.6) | 12,165 (13.8) |
| 2019 (May 2019 to April 2020) | 20,601 (11.7) | 8,311 (11.9) | 4,481 (11.4) | 5,172 (12.2) | 2,637 (10.5) | 16,333 (10.7) | 1,255 (14.4) | 2,093 (12.9) | 4,473 (11.5) | 8,512 (9.6) |
| 2020 (May to December) | 3,896 (2.2) | 2,583 (3.7) | 679 (1.7) | 475 (1.1) | 159 (0.6) | 411 (0.3) | 70 (0.8) | 78 (0.5) | 173 (0.4) | 90 (0.1) |
| 2021 (January to December) | 21,121 (12.0) | 7,858 (11.2) | 3,350 (8.5) | 5,363 (12.6) | 4,550 (18.2) | 1,101 (0.7.0) | 171 (2.0) | 160 (1.0) | 441 (1.1) | 329 (0.4) |
| 2022 (January to July) | 4,913 (2.8) | 1,946 (2.8) | 838 (2.1) | 1,105 (2.6) | 1,024 (4.1) | 948 (0.6) | 146 (1.7) | 128 (0.8) | 223 (0.6) | 451 (0.5) |
| **Comorbidity during baseline period, n (%)** | | | | | | | | | | |
| Down syndrome | 1,785 (1.0) | 975 (1.4) | 367 (0.9) | 262 (0.6) | 181 (0.7) | 189 (0.1) | 24 (0.3) | 55 (0.3) | 48 (0.1) | 62 (0.1) |
| Congenital heart disease | 9,150 (5.2) | 5,717 (8.2) | 2,065 (5.3) | 968 (2.3) | 400 (1.6) | 1,123 (0.7) | 231 (2.6) | 396 (2.4) | 277 (0.7) | 219 (0.3) |
| Bronchopulmonary dysplasia | 579 (0.3) | 228 (0.3) | 213 (0.5) | 96 (0.2) | 42 (0.2) | 57 (0.0) | 12 (0.1) | 17 (0.1) | 14 (0.0) | 14 (0.0) |
| Prematurity | 21,686 (12.3) | 17,297 (24.7) | 3,288 (8.4) | 760 (1.8) | 341 (1.4) | 851 (0.6) | 253 (2.9) | 456 (2.8) | 118 (0.3) | 24 (0.0) |
| Immunodeficiency | 795 (0.5) | 370 (0.5) | 201 (0.5) | 134 (0.3) | 90 (0.4) | 329 (0.2) | 39 (0.5) | 62 (0.4) | 75 (0.2) | 153 (0.2) |
| Pulmonary hypoplasia | 78 (0.0) | 43 (0.1) | 22 (0.1) | 9 (0.0) | 4 (0.0) | 13 (0.0) | 3 (0.0) | 7 (0.0) | 1 (0.0) | 2 (0.0) |
| Airway stenosis | 121 (0.1) | 30 (0.0) | 46 (0.1) | 32 (0.1) | 13 (0.1) | 31 (0.0) | 8 (0.1) | 8 (0.1) | 5 (0.0) | 10 (0.0) |
| Esophageal atresia | 6 (0.0) | 3 (0.0) | 0 (0.0) | 3 (0.0) | 0 (0.0) | 2 (0.0) | 0 (0.0) | 0 (0.0) | 1 (0.0) | 1 (0.0) |

| | | | | | | | | | | |
|---|---|---|---|---|---|---|---|---|---|---|
| Congenital metabolic disorder | 1,382 (0.8) | 492 (0.7) | 374 (1) | 311 (0.7) | 205 (0.8) | 780 (0.5) | 48 (0.6) | 108 (0.7) | 249 (0.6) | 375 (0.4) |
| Neuromuscular disorder | 1,078 (0.6) | 356 (0.5) | 243 (0.6) | 267 (0.6) | 212 (0.8) | 638 (0.4) | 27 (0.3) | 89 (0.6) | 220 (0.6) | 302 (0.3) |
| Cystic fibrosis | 0 (0.0) | 0 (0.0) | 0 (0.0) | 0 (0.0) | 0 (0.0) | 0 (0.0) | 0 (0.0) | 0 (0.0) | 0 (0.0) | 0 (0.0) |
| No record of these comorbidities | 144,611 (81.7) | 47,061 (67.2) | 33,654 (85.6) | 40,077 (94.3) | 23,819 (94.9) | 149,947 (97.8) | 8,203 (93.8) | 15,318 (94.0) | 38,329 (97.7) | 88,097 (98.8) |
| **Palivizumab use, n (%)** | | | | | | | | | | |
| Yes | 16,940 (9.6) | 12,356 (17.7) | 3,265 (8.3) | 1,172 (2.8) | 147 (0.6) | 1,131 (0.7) | 146 (1.7) | 404 (2.5) | 487 (1.2) | 94 (0.1) |
| No record of use | 159,971 (90.4) | 57,665 (82.4) | 36,046 (91.7) | 41,313 (97.2) | 24,947 (99.4) | 152,252 (99.3) | 8,595 (98.3) | 15,891 (97.5) | 38,731 (98.8) | 89,035 (99.9) |
| **Size of healthcare facilities, n (%)** | | | | | | | | | | |
| Less than 200 beds | 13,135 (7.4) | 4,296 (6.1) | 3,327 (8.5) | 3,462 (8.2) | 2,050 (8.2) | 14,403 (9.4) | 412 (4.7) | 1,310 (8.0) | 3,599 (9.2) | 9,082 (10.2) |
| 200 to 499 beds | 97,448 (55.1) | 35,523 (50.7) | 23,038 (58.6) | 24,579 (57.9) | 14,308 (57.0) | 95,370 (62.2) | 4,694 (53.7) | 9,971 (61.2) | 24,243 (61.8) | 56,462 (63.4) |
| 500 or more beds | 66,328 (37.5) | 30,202 (43.1) | 12,946 (32.9) | 14,444 (34.0) | 8,736 (34.8) | 43,610 (28.4) | 3,635 (41.6) | 5,014 (30.8) | 11,376 (29.0) | 23,585 (26.5) |

*Rounding may create totals slightly over or below 100% in some cases.*

**Supplementary Table 3** Baseline characteristics of healthy children <5 years of age who were diagnosed in the outpatient setting with RSV infection and influenza

| | RSV | | | | | Influenza | | | | |
|---|---|---|---|---|---|---|---|---|---|---|
| | **Total** | **Age <6 months** | **Age 6-11 months** | **Age 1<2 years** | **Age 2<5 years** | **Total** | **Age <6 months** | **Age 6-11 months** | **Age 1<2 years** | **Age 2<5 years** |
| **Total number of patients, n (relative % by age group)** | 141,843 (100) | 45,318 (31.9) | 33,225 (23.4) | 39,545 (27.9) | 23,755 (16.7) | 149,529 (100) | 8,197 (5.5) | 15,277 (10.2) | 38,019 (25.4) | 88,036 (58.9) |
| **Sex, n (%)** | | | | | | | | | | |
| Female | 63,794 (45.0) | 19,792 (43.7) | 14,462 (43.5) | 18,158 (45.9) | 11,382 (47.9) | 68,883 (46.1) | 3,643 (44.4) | 7,016 (45.9) | 17,600 (46.3) | 40,624 (46.1) |
| Male | 78,049 (55.0) | 25,526 (56.3) | 18,763 (56.5) | 21,387 (54.1) | 12,373 (52.1) | 80,646 (53.9) | 4,554 (55.6) | 8,261 (54.1) | 20,419 (53.7) | 47,412 (53.9) |
| **Age, months** | | | | | | | | | | |
| **Mean (SD)** | 13.3 (11.9) | 2.7 (1.4) | 8.4 (1.7) | 16.6 (3.4) | 35.0 (9.2) | 30.1 (16.7) | 3.1 (1.4) | 8.8 (1.7) | 17.1 (3.4) | 41.9 (10.5) |
| **Median (IQR)** | 10.0 (4.0-19.0) | 3.0 (2.0-4.0) | 8.0 (7.0-10.0) | 16.0 (14.0-19.0) | 33.0 (27.0-41.0) | 29.0 (15.0-45.0) | 3.0 (2.0-4.0) | 9.0 (7.0-10.0) | 17.0 (14.0-20.0) | 42.0 (33.0-51.0) |
| **Season by year, n (%)** | | | | | | | | | | |
| 2011 (May 2011 to April 2012) | 3,671 (2.6) | 1,063 (2.3) | 801 (2.4) | 1,101 (2.8) | 706 (3.0) | 8,198 (5.5) | 191 (2.3) | 571 (3.8) | 1,785 (4.7) | 5,651 (6.5) |
| 2012 (May 2012 to April 2013) | 6,880 (4.9) | 2,224 (4.9) | 1,722 (5.2) | 1,839 (4.7) | 1,095 (4.6) | 8,474 (5.7) | 300 (3.7) | 689 (4.5) | 1,957 (5.2) | 5,528 (6.3) |
| 2013 (May 2013 to April 2014) | 8,962 (6.3) | 3,161 (7.0) | 2,199 (6.6) | 2,290 (5.8) | 1,312 (5.5) | 13,133 (8.8) | 496 (6.1) | 1,133 (7.5) | 3,255 (8.6) | 8,249 (9.4) |
| 2014 (May 2014 to April 2015) | 13,808 (9.7) | 4,610 (10.2) | 3,385 (10.2) | 3,662 (9.3) | 2,151 (9.1) | 17,659 (11.9) | 860 (10.5) | 1,652 (10.9) | 4,452 (11.8) | 10,695 (12.2) |
| 2015 (May 2015 to April 2016) | 15,391 (10.9) | 5,114 (11.3) | 3,881 (11.7) | 4,128 (10.4) | 2,268 (9.6) | 19,475 (13.1) | 803 (9.8) | 1,923 (12.7) | 4,908 (13.0) | 11,841 (13.6) |
| 2016 (May 2016 to April 2017) | 14,698 (10.4) | 4,917 (10.9) | 3,672 (11.1) | 4,016 (10.2) | 2,093 (8.8) | 18,068 (12.2) | 1,032 (12.6) | 1,892 (12.4) | 4,819 (12.7) | 10,325 (11.8) |

| | | | | | | | | | | |
|---|---|---|---|---|---|---|---|---|---|---|
| 2017 (May 2017 to April 2018) | 21,058<br>(14.9) | 6,623<br>(14.6) | 5,240<br>(15.8) | 5,862<br>(14.8) | 3,333<br>(14.0) | 23,843<br>(16.0) | 1,389<br>(17.0) | 2,613<br>(17.2) | 5,999<br>(15.9) | 13,842<br>(15.8) |
| 2018 (May 2018 to April 2019) | 18,823<br>(13.3) | 6,395<br>(14.1) | 4,602<br>(13.9) | 5,174<br>(13.1) | 2,652<br>(11.2) | 21,540<br>(14.5) | 1,566<br>(19.1) | 2,431<br>(16.0) | 5,529<br>(14.6) | 12,014<br>(13.7) |
| 2019 (May 2019 to April 2020) | 16,484<br>(11.6) | 5,130<br>(11.3) | 3,916<br>(11.8) | 4,888<br>(12.4) | 2,550<br>(10.7) | 15,880<br>(10.7) | 1,182<br>(14.4) | 1,966<br>(12.9) | 4,322<br>(11.4) | 8,410<br>(9.6) |
| 2020 (May to December) | 1,162<br>(0.8) | 363<br>(0.8) | 280<br>(0.8) | 376<br>(1.0) | 143<br>(0.6) | 376<br>(0.3) | 59<br>(0.7) | 66<br>(0.4) | 168<br>(0.4) | 83<br>(0.1) |
| 2021 (January to December) | 17,211<br>(12.1) | 4,782<br>(10.6) | 2,868<br>(8.6) | 5,132<br>(13.0) | 4,429<br>(18.7) | 1,026<br>(0.7) | 162<br>(2.0) | 143<br>(0.9) | 413<br>(1.1) | 308<br>(0.4) |
| 2022 (January to July) | 3,587<br>(2.5) | 906<br>(2.0) | 638<br>(1.9) | 1,043<br>(2.6) | 1,000<br>(4.2) | 912<br>(0.6) | 140<br>(1.7) | 122<br>(0.8) | 214<br>(0.6) | 436<br>(0.5) |
| **Size of healthcare facilities, n (%)** | | | | | | | | | | |
| Less than 200 beds | 11,318<br>(8.0) | 3,069<br>(6.8) | 2,951<br>(8.9) | 3,312<br>(8.4) | 1,986<br>(8.4) | 14,273<br>(9.5) | 401<br>(4.9) | 1,281<br>(8.4) | 3,555<br>(9.4) | 9,036<br>(10.3) |
| 200 to 499 beds | 83,300<br>(58.7) | 25,757<br>(56.8) | 20,393<br>(61.4) | 23,359<br>(59.1) | 13,791<br>(58.1) | 93,555<br>(62.6) | 4,447<br>(54.3) | 9,467<br>(62.0) | 23,700<br>(62.3) | 55,941<br>(63.5) |
| 500 or more beds | 47,225<br>(33.3) | 16,492<br>(36.4) | 9,881<br>(29.7) | 12,874<br>(32.6) | 7,978<br>(33.6) | 41,701<br>(27.9) | 3,349<br>(40.9) | 4,529<br>(29.6) | 10,764<br>(28.3) | 23,059<br>(26.2) |

*Rounding may create totals slightly over or below 100% in some cases.*

**Supplementary Table 4.** Baseline characteristics for healthy children hospitalized with RSV infection and influenza overall and by age categories

| | RSV | | | | | Influenza | | | | |
|---|---|---|---|---|---|---|---|---|---|---|
| | Total | Age <6 months | Age 6-11 months | Age 1<2 years | Age 2<5 years | Total | Age <6 months | Age 6-11 months | Age 1<2 years | Age 2<5 years |
| **Total number of patients, n (relative % by age group)** | 85,550 (100) | 32,909 (38.5) | 17,618 (20.6) | 23,046 (26.9) | 11,977 (14.0) | 10,764 (100) | 2,064 (19.2) | 1,100 (10.2) | 2,728 (25.3) | 4,872 (45.3) |
| **Sex, n (%)** | | | | | | | | | | |
| Female | 38,766 (45.3) | 14,508 (44.1) | 7,489 (42.5) | 10,741 (46.6) | 6,028 (50.3) | 4,696 (43.6) | 952 (46.1) | 478 (43.5) | 1,228 (45.0) | 2,038 (41.8) |
| Male | 46,784 (54.7) | 18,401 (55.9) | 10,129 (57.5) | 12,305 (53.4) | 5,949 (49.7) | 6,068 (56.4) | 1,112 (53.9) | 622 (56.5) | 1,500 (55.0) | 2,834 (58.2) |
| **Age, months** | | | | | | | | | | |
| **Mean (SD)** | 11.9 (11.2) | 2.5 (1.4) | 8.4 (1.7) | 16.7 (3.4) | 33.9 (8.6) | 24.2 (17.4) | 2.5 (1.3) | 8.6 (1.7) | 17.4 (3.4) | 40.7 (10.6) |
| **Median (IQR)** | 9.0 (3.0-17.0) | 2.0 (1.0-4.0) | 8.0 (7.0-10.0) | 16.0 (14.0-19.0) | 31.0 (27.0-39.0) | 21.0 (9.0-38.5) | 2.0 (2.0-3.0) | 9.0 (7.0-10.0) | 17.0 (14.0-20.0) | 40.0 (31.0-50.0) |
| **Season by year, n (%)** | | | | | | | | | | |
| 2011 (May 2011 to April 2012) | 1,911 (2.2) | 684 (2.1) | 428 (2.4) | 530 (2.3) | 269 (2.2) | 353 (3.3) | 68 (3.3) | 36 (3.3) | 78 (2.9) | 171 (3.5) |
| 2012 (May 2012 to April 2013) | 3,448 (4.0) | 1,297 (3.9) | 799 (4.5) | 911 (4.0) | 441 (3.7) | 496 (4.6) | 97 (4.7) | 41 (3.7) | 124 (4.6) | 234 (4.8) |
| 2013 (May 2013 to April 2014) | 4,925 (5.8) | 2,105 (6.4) | 1,093 (6.2) | 1,169 (5.1) | 558 (4.7) | 900 (8.4) | 141 (6.8) | 91 (8.3) | 216 (7.9) | 452 (9.3) |
| 2014 (May 2014 to April 2015) | 7,587 (8.9) | 2,987 (9.1) | 1,684 (9.6) | 1,961 (8.5) | 955 (8.0) | 1,088 (10.1) | 251 (12.2) | 107 (9.8) | 254 (9.3) | 476 (9.8) |
| 2015 (May 2015 to April 2016) | 8,975 (10.5) | 3,565 (10.8) | 1,923 (10.9) | 2,366 (10.3) | 1,121 (9.4) | 1,507 (14.0) | 225 (10.9) | 188 (17.1) | 384 (14.1) | 710 (14.6) |
| 2016 (May 2016 to April 2017) | 8,375 (9.8) | 3,343 (10.2) | 1,906 (10.8) | 2,214 (9.6) | 912 (7.6) | 1,334 (12.4) | 293 (14.2) | 119 (10.8) | 337 (12.4) | 585 (12.1) |

| | | | | | | | | | | |
|---|---|---|---|---|---|---|---|---|---|---|
| 2017 (May 2017 to April 2018) | 12,640<br>(14.8) | 4,763<br>(14.5) | 2,798<br>(15.9) | 3,436<br>(14.9) | 1,643<br>(13.7) | 1,612<br>(15.0) | 292<br>(14.2) | 162<br>(14.8) | 381<br>(14.0) | 777<br>(16.0) |
| 2018 (May 2018 to April 2019) | 11,639<br>(13.6) | 4,739<br>(14.4) | 2,556<br>(14.5) | 3,034<br>(13.2) | 1,310<br>(10.9) | 1,818<br>(16.9) | 401<br>(19.5) | 173<br>(15.8) | 469<br>(17.2) | 775<br>(16.0) |
| 2019 (May 2019 to April 2020) | 11,031<br>(12.9) | 4,107<br>(12.5) | 2,252<br>(12.8) | 3,217<br>(14.0) | 1,455<br>(12.2) | 1,608<br>(15.0) | 288<br>(14.0) | 177<br>(16.1) | 474<br>(17.4) | 669<br>(13.8) |
| 2020 (May to December) | 711<br>(0.8) | 239<br>(0.7) | 151<br>(0.9) | 231<br>(1.0) | 90<br>(0.8) | 3<br>(0.0) | 1<br>(0.0) | 2<br>(0.2) | 0<br>(0.0) | 0<br>(0.0) |
| 2021 (January to December) | 11,924<br>(13.9) | 4,261<br>(13.0) | 1,671<br>(9.5) | 3,341<br>(14.5) | 2,651<br>(22.1) | 6<br>(0.1) | 1<br>(0.0) | 1<br>(0.1) | 3<br>(0.1) | 1<br>(0.0) |
| 2022 (January to July) | 2,328<br>(2.7) | 797<br>(2.4) | 346<br>(2.0) | 619<br>(2.7) | 566<br>(4.7) | 4<br>(0.0) | 1<br>(0.0) | 0<br>(0.0) | 3<br>(0.1) | 0<br>(0.0) |
| **Size of healthcare facilities, n (%)** | | | | | | | | | | |
| Less than 200 beds | 5,072<br>(5.9) | 1,700<br>(5.2) | 1,165<br>(6.6) | 1,420<br>(6.2) | 787<br>(6.6) | 541<br>(5.0) | 66<br>(3.2) | 54<br>(4.9) | 152<br>(5.6) | 269<br>(5.5) |
| 200 to 499 beds | 48,569<br>(56.8) | 17,932<br>(54.5) | 10,427<br>(59.2) | 13,476<br>(58.5) | 6,734<br>(56.2) | 5,907<br>(54.9) | 1,100<br>(53.3) | 597<br>(54.3) | 1,529<br>(56.0) | 2,681<br>(55.0) |
| 500 or more beds | 31,909<br>(37.3) | 13,277<br>(40.3) | 6,026<br>(34.2) | 8,150<br>(35.4) | 4,456<br>(37.2) | 4,316<br>(40.1) | 898<br>(43.5) | 449<br>(40.8) | 1,047<br>(38.4) | 1,922<br>(39.4) |

*Rounding may create totals slightly over or below 100% in some cases.*

**Supplementary Table 5.** Distribution of infants <1 year of age who are diagnosed in the outpatient setting, who are hospitalized, who required oxygen use, or who required mechanical ventilation with RSV infection or influenza in Japan by month of age

| Age (month) | 0 | 1 | 2 | 3 | 4 | 5 | 6 | 7 | 8 | 9 | 10 | 11 |
|---|---|---|---|---|---|---|---|---|---|---|---|---|
| | n (%) | n (%) | n (%) | n (%) | n (%) | n (%) | n (%) | n (%) | n (%) | n (%) | n (%) | n (%) |
| **RSV infection** | | | | | | | | | | | | |
| **All children: Total number of patient who are diagnosed in the outpatient setting per age group, n (%)** | 4,311 (3.9) | 15,885 (14.5) | 16,961 (15.5) | 12,944 (11.8) | 10,884 (10.0) | 9,036 (8.3) | 7,830 (7.2) | 7,026 (6.4) | 6,311 (5.8) | 6,149 (5.6) | 5,967 (5.5) | 6,028 (5.5) |
| **All children: Total number of hospitalized patients per age group, n (%)** | 1,271 (2.3) | 8,223 (15.2) | 9,500 (17.6) | 6,901 (12.8) | 5,230 (9.7) | 4,148 (7.7) | 3,546 (6.6) | 3,362 (6.2) | 2,999 (5.5) | 3,023 (5.6) | 2,940 (5.4) | 2,957 (5.5) |
| **All children: Total number of patient who required mechanical ventilation per age group, n (%)** | 71 (4.8) | 454 (30.8) | 326 (22.1) | 131 (8.9) | 94 (6.4) | 79 (5.4) | 55 (3.7) | 60 (4.1) | 52 (3.5) | 54 (3.7) | 50 (3.4) | 46 (3.1) |
| **Healthy children: Total number of patient who are diagnosed in the outpatient setting per age group, n (%)** | 1,379 (1.8%) | 9,044 (11.5%) | 11,192 (14.2%) | 9,200 (11.7%) | 7,849 (10%) | 6,654 (8.5%) | 5,999 (7.6%) | 5,759 (7.3%) | 5,389 (6.9%) | 5,343 (6.8%) | 5,339 (6.8%) | 5,396 (6.9%) |
| **Healthy children: Total number of hospitalized patients per age group, n (%)** | 1,134 (2.2%) | 7,691 (15.2%) | 8,894 (17.6%) | 6,500 (12.9%) | 4,849 (9.6%) | 3,841 (7.6%) | 3,269 (6.5%) | 3,152 (6.2%) | 2,801 (5.5%) | 2,819 (5.6%) | 2,801 (5.5%) | 2,776 (5.5%) |
| **Healthy children: Total number of patient who required mechanical ventilation per age group, n (%)** | 62 (4.8) | 419 (32.2) | 291 (22.4) | 121 (9.3) | 83 (6.4) | 57 (4.4) | 45 (3.5) | 50 (3.8) | 42 (3.2) | 46 (3.5) | 44 (3.4) | 42 (3.2) |
| **All Children: Overall hospitalizations after diagnosis (n, % of outpatient diagnoses)** | | | | | | | | | | | | |
| Admissions within 0-6 days | 570 (13.2) | 3,884 (24.5) | 4,477 (26.4) | 3,245 (25.1) | 2,449 (22.5) | 1,935 (21.4) | 1,659 (21.2) | 1,606 (22.9) | 1,416 (22.4) | 1,434 (23.3) | 1,381 (23.1) | 1,407 (23.3) |
| Admissions within 0-13 days | 570 | 3,912 | 4,496 | 3,258 | 2,460 | 1,949 | 1,672 | 1,613 | 1,423 | 1,446 | 1,386 | 1,409 |

| | | | | | | | | | | | | |
|---|---|---|---|---|---|---|---|---|---|---|---|---|
| | (13.2) | (24.6) | (26.5) | (25.2) | (22.6) | (21.6) | (21.4) | (23.0) | (22.5) | (23.5) | (23.2) | (23.4) |
| Admissions within 0-29 days | 576 | 3,952 | 4,561 | 3,306 | 2,495 | 1,979 | 1,694 | 1,641 | 1,448 | 1,465 | 1,410 | 1,432 |
| | (13.4) | (24.9) | (26.9) | (25.5) | (22.9) | (21.9) | (21.6) | (23.4) | (22.9) | (23.8) | (23.6) | (23.8) |
| **Healthy Children: Overall hospitalizations after diagnosis (n, % of outpatient diagnoses)** | | | | | | | | | | | | |
| Admissions within 0-6 days | 541 | 3,699 | 4,267 | 3,100 | 2,297 | 1,813 | 1,562 | 1,541 | 1,338 | 1,346 | 1,323 | 1,333 |
| | (39.2) | (40.9) | (38.1) | (33.7) | (29.3) | (27.2) | (26.0) | (26.8) | (24.8) | (25.2) | (24.8) | (24.7) |
| Admissions within 0-13 days | 541 | 3,726 | 4,283 | 3,112 | 2,306 | 1,826 | 1,573 | 1,547 | 1,345 | 1,356 | 1,325 | 1,333 |
| | (39.2) | (41.2) | (38.3) | (33.8) | (29.4) | (27.4) | (26.2) | (26.9) | (25.0) | (25.4) | (24.8) | (24.7) |
| Admissions within 0-29 days | 545 | 3,754 | 4,331 | 3,156 | 2,336 | 1,848 | 1,593 | 1,573 | 1,368 | 1,374 | 1,348 | 1,354 |
| | (39.5) | (41.5) | (38.7) | (34.3) | (29.8) | (27.8) | (26.6) | (27.3) | (25.4) | (25.7) | (25.2) | (25.1) |
| **Influenza** | | | | | | | | | | | | |
| **All children: Total number of patient who are diagnosed in the outpatient setting per age group, n (%)** | 215 | 1,178 | 1,823 | 1,908 | 1,816 | 1,801 | 2,088 | 2,315 | 2,604 | 2,934 | 3,121 | 3,233 |
| | (0.9) | (4.7) | (7.3) | (7.6) | (7.3) | (7.2) | (8.3) | (9.2) | (10.4) | (11.7) | (12.5) | (12.9) |
| **All children: Total number of hospitalized patients per age group, n (%)** | 53 | 405 | 655 | 519 | 327 | 182 | 193 | 168 | 184 | 206 | 222 | 215 |
| | (1.6) | (12.2) | (19.7) | (15.6) | (9.8) | (5.5) | (5.8) | (5.0) | (5.5) | (6.2) | (6.7) | (6.5) |
| **All children: Total number of patient who required mechanical ventilation per age group, n (%)** | 0 | 1 | 4 | 0 | 2 | 1 | 0 | 2 | 0 | 1 | 1 | 1 |
| | (0.0) | (7.7) | (30.8) | (0.0) | (15.4) | (7.7) | (0.0) | (15.4) | (0.0) | (7.7) | (7.7) | (7.7) |
| **Healthy children: Total number of patient who are diagnosed in the outpatient setting per age group, n (%)** | 192 | 1,086 | 1,723 | 1,810 | 1,708 | 1,678 | 1,949 | 2,174 | 2,443 | 2,739 | 2,944 | 3,028 |
| | (0.8%) | (4.6%) | (7.3%) | (7.7%) | (7.3%) | (7.1%) | (8.3%) | (9.3%) | (10.4%) | (11.7%) | (12.5%) | (12.9%) |
| **Healthy children: Total number of hospitalized patients per age group, n (%)** | 51 | 394 | 637 | 500 | 315 | 167 | 183 | 153 | 174 | 189 | 202 | 199 |
| | (1.6%) | (12.5%) | (20.1%) | (15.8%) | (10%) | (5.3%) | (5.8%) | (4.8%) | (5.5%) | (6%) | (6.4%) | (6.3%) |
| **Healthy children: Total number of patient who required mechanical ventilation per age group, n (%)** | 0 | 1 | 3 | 0 | 0 | 0 | 0 | 1 | 0 | 1 | 1 | 0 |
| | (0%) | (14.3%) | (42.9%) | (0%) | (0%) | (0%) | (0%) | (14.3%) | (0%) | (14.3%) | (14.3%) | (0%) |

| | | | | | | | | | | | | |
|---|---|---|---|---|---|---|---|---|---|---|---|---|
| **All Children: Overall hospitalizations after diagnosis (n, % of outpatient diagnoses)** | | | | | | | | | | | | |
| Admissions within 0-6 days | 17 | 128 | 216 | 174 | 124 | 77 | 66 | 56 | 65 | 82 | 90 | 82 |
| | (7.9) | (10.9) | (11.8) | (9.1) | (6.8) | (4.3) | (3.2) | (2.4) | (2.5) | (2.8) | (2.9) | (2.5) |
| Admissions within 0-13 days | 17 | 129 | 217 | 174 | 125 | 77 | 66 | 56 | 65 | 82 | 90 | 85 |
| | (7.9) | (11.0) | (11.9) | (9.1) | (6.9) | (4.3) | (3.2) | (2.4) | (2.5) | (2.8) | (2.9) | (2.6) |
| Admissions within 0-29 days | 17 | 129 | 217 | 174 | 126 | 77 | 67 | 56 | 65 | 82 | 92 | 86 |
| | (7.9) | (11.0) | (11.9) | (9.1) | (6.9) | (4.3) | (3.2) | (2.4) | (2.5) | (2.8) | (2.9) | (2.7) |
| **Healthy Children: Overall hospitalizations after diagnosis (n, % of outpatient diagnoses)** | | | | | | | | | | | | |
| Admissions within 0-6 days | 16 | 121 | 206 | 163 | 119 | 69 | 61 | 47 | 58 | 70 | 76 | 74 |
| | (8.3) | (11.1) | (12.0) | (9.0) | (7.0) | (4.1) | (3.1) | (2.2) | (2.4) | (2.6) | (2.6) | (2.4) |
| Admissions within 0-13 days | 16 | 122 | 207 | 163 | 120 | 69 | 61 | 47 | 58 | 70 | 76 | 77 |
| | (8.3) | (11.2) | (12.0) | (9.0) | (7.0) | (4.1) | (3.1) | (2.2) | (2.4) | (2.6) | (2.6) | (2.5) |
| Admissions within 0-29 days | 16 | 122 | 207 | 163 | 121 | 69 | 62 | 47 | 58 | 70 | 77 | 78 |
| | (8.3) | (11.2) | (12.0) | (9.0) | (7.1) | (4.1) | (3.2) | (2.2) | (2.4) | (2.6) | (2.6) | (2.6) |

*Rounding may create totals slightly over or below 100% in some cases.*

**Supplementary Table 6.** Annual number of cases by month

| Year | Month | RSV - Inpatients | | Influenza - Inpatients | | RSV - Outpatients | | Influenza - Outpatients | |
|---|---|---|---|---|---|---|---|---|---|
| | | n | % | n | % | n | % | n | % |
| 2011 | 4 | 56 | 0.1 | 36 | 0.3 | 114 | 0.1 | 954 | 0.6 |
| | 5 | 44 | 0.0 | 22 | 0.2 | 92 | 0.1 | 379 | 0.2 |
| | 6 | 60 | 0.1 | 6 | 0.1 | 103 | 0.1 | 184 | 0.1 |
| | 7 | 77 | 0.1 | 1 | 0.0 | 145 | 0.1 | 50 | 0.0 |
| | 8 | 127 | 0.1 | 0 | 0.0 | 231 | 0.1 | 17 | 0.0 |
| | 9 | 178 | 0.2 | 1 | 0.0 | 398 | 0.2 | 23 | 0.0 |
| | 10 | 219 | 0.2 | 0 | 0.0 | 466 | 0.3 | 54 | 0.0 |
| | 11 | 267 | 0.3 | 9 | 0.1 | 596 | 0.3 | 164 | 0.1 |
| | 12 | 451 | 0.5 | 19 | 0.2 | 845 | 0.5 | 578 | 0.4 |
| 2012 | 1 | 252 | 0.3 | 74 | 0.7 | 550 | 0.3 | 1,937 | 1.3 |
| | 2 | 124 | 0.1 | 111 | 1.0 | 276 | 0.2 | 2,752 | 1.8 |
| | 3 | 129 | 0.1 | 77 | 0.7 | 287 | 0.2 | 1,423 | 0.9 |
| | 4 | 72 | 0.1 | 40 | 0.4 | 202 | 0.1 | 747 | 0.5 |
| | 5 | 52 | 0.1 | 11 | 0.1 | 117 | 0.1 | 236 | 0.2 |
| | 6 | 80 | 0.1 | 6 | 0.1 | 164 | 0.1 | 99 | 0.1 |
| | 7 | 83 | 0.1 | 4 | 0.0 | 183 | 0.1 | 114 | 0.1 |
| | 8 | 206 | 0.2 | 6 | 0.1 | 273 | 0.2 | 91 | 0.1 |
| | 9 | 354 | 0.4 | 0 | 0.0 | 1,139 | 0.6 | 74 | 0.0 |
| | 10 | 596 | 0.7 | 1 | 0.0 | 1,430 | 0.8 | 66 | 0.0 |
| | 11 | 599 | 0.7 | 4 | 0.0 | 1,291 | 0.7 | 133 | 0.1 |
| | 12 | 699 | 0.8 | 22 | 0.2 | 1,430 | 0.8 | 499 | 0.3 |
| 2013 | 1 | 401 | 0.4 | 177 | 1.6 | 887 | 0.5 | 2,753 | 1.8 |
| | 2 | 188 | 0.2 | 154 | 1.4 | 488 | 0.3 | 2,524 | 1.6 |
| | 3 | 191 | 0.2 | 84 | 0.8 | 461 | 0.3 | 1,275 | 0.8 |
| | 4 | 160 | 0.2 | 48 | 0.4 | 376 | 0.2 | 755 | 0.5 |
| | 5 | 132 | 0.1 | 37 | 0.3 | 297 | 0.2 | 626 | 0.4 |
| | 6 | 116 | 0.1 | 7 | 0.1 | 248 | 0.1 | 139 | 0.1 |
| | 7 | 199 | 0.2 | 1 | 0.0 | 332 | 0.2 | 58 | 0.0 |
| | 8 | 372 | 0.4 | 0 | 0.0 | 660 | 0.4 | 22 | 0.0 |
| | 9 | 651 | 0.7 | 1 | 0.0 | 1,621 | 0.9 | 19 | 0.0 |
| | 10 | 743 | 0.8 | 1 | 0.0 | 1,603 | 0.9 | 62 | 0.0 |
| | 11 | 850 | 0.9 | 8 | 0.1 | 1,733 | 1.0 | 229 | 0.1 |
| | 12 | 894 | 1.0 | 36 | 0.3 | 1,663 | 0.9 | 652 | 0.4 |
| 2014 | 1 | 542 | 0.6 | 222 | 2.0 | 1,142 | 0.6 | 2,746 | 1.8 |
| | 2 | 285 | 0.3 | 284 | 2.5 | 679 | 0.4 | 3,870 | 2.5 |
| | 3 | 232 | 0.3 | 212 | 1.9 | 607 | 0.3 | 2,806 | 1.8 |
| | 4 | 243 | 0.3 | 124 | 1.1 | 700 | 0.4 | 2,203 | 1.4 |
| | 5 | 204 | 0.2 | 43 | 0.4 | 540 | 0.3 | 846 | 0.6 |

| Year | Month | RSV - Inpatients | | Influenza - Inpatients | | RSV - Outpatients | | Influenza - Outpatients | |
|---|---|---|---|---|---|---|---|---|---|
| | | n | % | n | % | n | % | n | % |
| | 6 | 176 | 0.2 | 9 | 0.1 | 422 | 0.2 | 240 | 0.2 |
| | 7 | 225 | 0.2 | 2 | 0.0 | 472 | 0.3 | 126 | 0.1 |
| | 8 | 379 | 0.4 | 4 | 0.0 | 689 | 0.4 | 50 | 0.0 |
| | 9 | 850 | 0.9 | 1 | 0.0 | 2,206 | 1.2 | 93 | 0.1 |
| | 10 | 937 | 1.0 | 3 | 0.0 | 1,933 | 1.1 | 140 | 0.1 |
| | 11 | 1,405 | 1.6 | 26 | 0.2 | 2,542 | 1.4 | 504 | 0.3 |
| | 12 | 1,781 | 2.0 | 210 | 1.9 | 3,307 | 1.9 | 3,406 | 2.2 |
| | 1 | 794 | 0.9 | 545 | 4.9 | 1,653 | 0.9 | 8,043 | 5.2 |
| | 2 | 467 | 0.5 | 162 | 1.4 | 1,025 | 0.6 | 2,653 | 1.7 |
| | 3 | 433 | 0.5 | 87 | 0.8 | 1,060 | 0.6 | 1,133 | 0.7 |
| | 4 | 406 | 0.4 | 50 | 0.4 | 1,060 | 0.6 | 850 | 0.6 |
| | 5 | 218 | 0.2 | 17 | 0.2 | 593 | 0.3 | 421 | 0.3 |
| | 6 | 240 | 0.3 | 7 | 0.1 | 523 | 0.3 | 178 | 0.1 |
| 2015 | 7 | 336 | 0.4 | 4 | 0.0 | 530 | 0.3 | 120 | 0.1 |
| | 8 | 555 | 0.6 | 2 | 0.0 | 913 | 0.5 | 82 | 0.1 |
| | 9 | 905 | 1.0 | 6 | 0.1 | 2,333 | 1.3 | 116 | 0.1 |
| | 10 | 1,322 | 1.5 | 10 | 0.1 | 2,658 | 1.5 | 257 | 0.2 |
| | 11 | 1,922 | 2.1 | 22 | 0.2 | 3,442 | 1.9 | 462 | 0.3 |
| | 12 | 2,070 | 2.3 | 43 | 0.4 | 3,611 | 2.0 | 868 | 0.6 |
| | 1 | 874 | 1.0 | 222 | 2.0 | 1,676 | 0.9 | 3,103 | 2.0 |
| | 2 | 401 | 0.4 | 689 | 6.2 | 943 | 0.5 | 7,554 | 4.9 |
| | 3 | 338 | 0.4 | 395 | 3.5 | 852 | 0.5 | 4,625 | 3.0 |
| | 4 | 351 | 0.4 | 158 | 1.4 | 779 | 0.4 | 2,174 | 1.4 |
| | 5 | 287 | 0.3 | 34 | 0.3 | 671 | 0.4 | 610 | 0.4 |
| 2016 | 6 | 255 | 0.3 | 4 | 0.0 | 584 | 0.3 | 141 | 0.1 |
| | 7 | 475 | 0.5 | 7 | 0.1 | 838 | 0.5 | 90 | 0.1 |
| | 8 | 671 | 0.7 | 3 | 0.0 | 1,129 | 0.6 | 75 | 0.0 |
| | 9 | 1,400 | 1.5 | 4 | 0.0 | 3,197 | 1.8 | 119 | 0.1 |
| | 10 | 1,848 | 2.0 | 24 | 0.2 | 3,456 | 2.0 | 485 | 0.3 |
| | 11 | 1,311 | 1.5 | 45 | 0.4 | 2,376 | 1.3 | 859 | 0.6 |
| | 12 | 997 | 1.1 | 133 | 1.2 | 1,906 | 1.1 | 2,094 | 1.4 |
| | 1 | 518 | 0.6 | 408 | 3.6 | 1,187 | 0.7 | 4,963 | 3.2 |
| | 2 | 299 | 0.3 | 375 | 3.4 | 820 | 0.5 | 4,666 | 3.0 |
| | 3 | 381 | 0.4 | 205 | 1.8 | 951 | 0.5 | 2,478 | 1.6 |
| 2017 | 4 | 484 | 0.5 | 136 | 1.2 | 1,127 | 0.6 | 1,980 | 1.3 |
| | 5 | 509 | 0.6 | 55 | 0.5 | 1,046 | 0.6 | 1,038 | 0.7 |
| | 6 | 557 | 0.6 | 16 | 0.1 | 1,004 | 0.6 | 351 | 0.2 |
| | 7 | 1,071 | 1.2 | 11 | 0.1 | 1,767 | 1.0 | 319 | 0.2 |
| | 8 | 2,434 | 2.7 | 20 | 0.2 | 4,025 | 2.3 | 356 | 0.2 |

| Year | Month | RSV - Inpatients | | Influenza - Inpatients | | RSV - Outpatients | | Influenza - Outpatients | |
|---|---|---|---|---|---|---|---|---|---|
| | | n | % | n | % | n | % | n | % |
| | 9 | 2,781 | 3.1 | 12 | 0.1 | 5,531 | 3.1 | 431 | 0.3 |
| | 10 | 1,692 | 1.9 | 27 | 0.2 | 3,154 | 1.8 | 491 | 0.3 |
| | 11 | 1,031 | 1.1 | 53 | 0.5 | 1,979 | 1.1 | 825 | 0.5 |
| | 12 | 976 | 1.1 | 247 | 2.2 | 1,818 | 1.0 | 3,546 | 2.3 |
| 2018 | 1 | 675 | 0.7 | 600 | 5.4 | 1,464 | 0.8 | 8,322 | 5.4 |
| | 2 | 490 | 0.5 | 404 | 3.6 | 1,118 | 0.6 | 5,553 | 3.6 |
| | 3 | 581 | 0.6 | 159 | 1.4 | 1,210 | 0.7 | 2,203 | 1.4 |
| | 4 | 637 | 0.7 | 73 | 0.7 | 1,288 | 0.7 | 1,109 | 0.7 |
| | 5 | 627 | 0.7 | 21 | 0.2 | 1,217 | 0.7 | 391 | 0.3 |
| | 6 | 657 | 0.7 | 14 | 0.1 | 1,239 | 0.7 | 227 | 0.1 |
| | 7 | 1,364 | 1.5 | 8 | 0.1 | 2,245 | 1.3 | 190 | 0.1 |
| | 8 | 2,031 | 2.2 | 8 | 0.1 | 3,787 | 2.1 | 161 | 0.1 |
| | 9 | 2,166 | 2.4 | 6 | 0.1 | 3,989 | 2.3 | 235 | 0.2 |
| | 10 | 1,352 | 1.5 | 14 | 0.1 | 2,546 | 1.4 | 358 | 0.2 |
| | 11 | 732 | 0.8 | 28 | 0.3 | 1,485 | 0.8 | 490 | 0.3 |
| | 12 | 819 | 0.9 | 215 | 1.9 | 1,526 | 0.9 | 2,432 | 1.6 |
| 2019 | 1 | 584 | 0.6 | 1,086 | 9.7 | 1,274 | 0.7 | 11,605 | 7.6 |
| | 2 | 530 | 0.6 | 353 | 3.2 | 1,036 | 0.6 | 3,923 | 2.6 |
| | 3 | 673 | 0.7 | 74 | 0.7 | 1,265 | 0.7 | 1,093 | 0.7 |
| | 4 | 821 | 0.9 | 68 | 0.6 | 1,534 | 0.9 | 1,024 | 0.7 |
| | 5 | 597 | 0.7 | 36 | 0.3 | 1,177 | 0.7 | 706 | 0.5 |
| | 6 | 528 | 0.6 | 13 | 0.1 | 975 | 0.6 | 295 | 0.2 |
| | 7 | 1,386 | 1.5 | 12 | 0.1 | 2,418 | 1.4 | 317 | 0.2 |
| | 8 | 2,328 | 2.6 | 13 | 0.1 | 3,826 | 2.2 | 348 | 0.2 |
| | 9 | 2,671 | 3.0 | 51 | 0.5 | 4,241 | 2.4 | 953 | 0.6 |
| | 10 | 1,492 | 1.7 | 48 | 0.4 | 2,493 | 1.4 | 707 | 0.5 |
| | 11 | 731 | 0.8 | 92 | 0.8 | 1,384 | 0.8 | 915 | 0.6 |
| | 12 | 662 | 0.7 | 547 | 4.9 | 1,322 | 0.7 | 4,537 | 3.0 |
| 2020 | 1 | 410 | 0.5 | 663 | 5.9 | 971 | 0.5 | 5,536 | 3.6 |
| | 2 | 355 | 0.4 | 153 | 1.4 | 799 | 0.5 | 1,452 | 0.9 |
| | 3 | 276 | 0.3 | 26 | 0.2 | 634 | 0.4 | 431 | 0.3 |
| | 4 | 140 | 0.2 | 6 | 0.1 | 361 | 0.2 | 136 | 0.1 |
| | 5 | 27 | 0.0 | 1 | 0.0 | 112 | 0.1 | 33 | 0.0 |
| | 6 | 29 | 0.0 | 0 | 0.0 | 231 | 0.1 | 33 | 0.0 |
| | 7 | 68 | 0.1 | 0 | 0.0 | 697 | 0.4 | 27 | 0.0 |
| | 8 | 59 | 0.1 | 0 | 0.0 | 649 | 0.4 | 22 | 0.0 |
| | 9 | 106 | 0.1 | 1 | 0.0 | 592 | 0.3 | 29 | 0.0 |
| | 10 | 159 | 0.2 | 0 | 0.0 | 607 | 0.3 | 53 | 0.0 |
| | 11 | 172 | 0.2 | 0 | 0.0 | 521 | 0.3 | 91 | 0.1 |

| Year | Month | RSV - Inpatients | | Influenza - Inpatients | | RSV - Outpatients | | Influenza - Outpatients | |
|---|---|---|---|---|---|---|---|---|---|
| | | n | % | n | % | n | % | n | % |
| 2021 | 12 | 182 | 0.2 | 1 | 0.0 | 487 | 0.3 | 123 | 0.1 |
| | 1 | 163 | 0.2 | 1 | 0.0 | 434 | 0.2 | 104 | 0.1 |
| | 2 | 228 | 0.3 | 1 | 0.0 | 417 | 0.2 | 88 | 0.1 |
| | 3 | 465 | 0.5 | 1 | 0.0 | 621 | 0.4 | 106 | 0.1 |
| | 4 | 933 | 1.0 | 1 | 0.0 | 1,265 | 0.7 | 137 | 0.1 |
| | 5 | 1,537 | 1.7 | 0 | 0.0 | 2,163 | 1.2 | 141 | 0.1 |
| | 6 | 2,348 | 2.6 | 0 | 0.0 | 3,881 | 2.2 | 144 | 0.1 |
| | 7 | 3,135 | 3.5 | 0 | 0.0 | 5,560 | 3.1 | 108 | 0.1 |
| | 8 | 1,884 | 2.1 | 0 | 0.0 | 3,451 | 2.0 | 65 | 0.0 |
| | 9 | 772 | 0.9 | 0 | 0.0 | 1,369 | 0.8 | 37 | 0.0 |
| | 10 | 326 | 0.4 | 0 | 0.0 | 726 | 0.4 | 29 | 0.0 |
| | 11 | 228 | 0.3 | 1 | 0.0 | 563 | 0.3 | 47 | 0.0 |
| | 12 | 377 | 0.4 | 1 | 0.0 | 671 | 0.4 | 95 | 0.1 |
| 2022 | 1 | 330 | 0.4 | 2 | 0.0 | 588 | 0.3 | 184 | 0.1 |
| | 2 | 160 | 0.2 | 0 | 0.0 | 373 | 0.2 | 149 | 0.1 |
| | 3 | 145 | 0.2 | 0 | 0.0 | 280 | 0.2 | 132 | 0.1 |
| | 4 | 81 | 0.1 | 0 | 0.0 | 213 | 0.1 | 108 | 0.1 |
| | 5 | 155 | 0.2 | 0 | 0.0 | 437 | 0.2 | 82 | 0.1 |
| | 6 | 404 | 0.4 | 0 | 0.0 | 745 | 0.4 | 60 | 0.0 |
| | 7 | 1,135 | 1.3 | 2 | 0.0 | 2,277 | 1.3 | 233 | 0.2 |

**Supplementary Table 7.** Baseline characteristics of children <5 years of age who are hospitalized with RSV infection by age group and COVID-19 era

| | Age <6 months | | | Age 6-11 months | | | Age 1-<2- years | | | Age 2-<5 years | | |
|---|---|---|---|---|---|---|---|---|---|---|---|---|
| | **Pre-COVID-19** | **During COVID-19** | **2021 season** | **Pre-COVID-19** | **During COVID-19** | **2021 season** | **Pre-COVID-19** | **During COVID-19** | **2021 season** | **Pre-COVID-19** | **During COVID-19** | **2021 season** |
| **Total number of patients per age group per time period, n (relative % by age group)** | 14,877 (39.3) | 6,106 (36.4) | 4,456 (35.9) | 8,311 (21.9) | 2,536 (15.1) | 1,771 (14.3) | 10,142 (26.8) | 4,574 (27.2) | 3,438 (27.7) | 4,537 (12.0) | 3,573 (21.3) | 2,731 (22.0) |
| **Sex, n (%)** | | | | | | | | | | | | |
| Female | 6,501 (43.7) | 2,760 (45.2) | 2,023 (45.4) | 3,518 (42.3) | 1,067 (42.1) | 732 (41.3) | 4,775 (47.1) | 2,074 (45.3) | 1,582 (46.0) | 2,228 (49.1) | 1,813 (50.7) | 1,396 (51.1) |
| Male | 8,376 (56.3) | 3,346 (54.8) | 2,433 (54.6) | 4,793 (57.7) | 1,469 (57.9) | 1,039 (58.7) | 5,367 (52.9) | 2,500 (54.7) | 1,856 (54.0) | 2,309 (50.9) | 1,760 (49.3) | 1,335 (48.9) |
| **Age, months** | | | | | | | | | | | | |
| **Mean (SD)** | 2.6 (1.4) | 2.4 (1.4) | 2.3 (1.4) | 8.4 (1.7) | 8.5 (1.7) | 8.4 (1.7) | 16.5 (3.3) | 17.3 (3.4) | 17.4 (3.4) | 33.7 (8.5) | 34.0 (8.8) | 33.8 (8.7) |
| **Median (IQR)** | 2.0 (1.0-4.0) | 2.0 (1.0-3.0) | 2.0 (1.0-3.0) | 8.0 (7.0-10.0) | 8.0 (7.0-10.0) | 8.0 (7.0-10.0) | 16.0 (14.0-19.0) | 17.0 (14.0-20.0) | 17.0 (15.0-20.0) | 31.0 (27.0-39.0) | 31.0 (27.0-39.0) | 31.0 (27.0-39.0) |
| **Season by year, n (%)** | | | | | | | | | | | | |
| 2016 (May 2016 to April 2017) | 758 (5.1) | 0 (0.0) | 0 (0.0) | 425 (5.1) | 0 (0.0) | 0 (0.0) | 353 (3.5) | 0 (0.0) | 0 (0.0) | 146 (3.2) | 0 (0.0) | 0 (0.0) |
| 2017 (May 2017 to April 2018) | 5,148 (34.6) | 0 (0.0) | 0 (0.0) | 2,987 (35.9) | 0 (0.0) | 0 (0.0) | 3,588 (35.4) | 0 (0.0) | 0 (0.0) | 1,711 (37.7) | 0 (0.0) | 0 (0.0) |
| 2018 (May 2018 to April 2019) | 5,105 (34.3) | 0 (0.0) | 0 (0.0) | 2,754 (33.1) | 0 (0.0) | 0 (0.0) | 3,148 (31.0) | 0 (0.0) | 0 (0.0) | 1,349 (29.7) | 0 (0.0) | 0 (0.0) |
| 2019 (May 2019 to April 2020) | 3,866 (26.0) | 514 (8.4) | 0 (0.0) | 2,145 (25.8) | 237 (9.3) | 0 (0.0) | 3,053 (30.1) | 260 (5.7) | 0 (0.0) | 1,331 (29.3) | 170 (4.8) | 0 (0.0) |
| 2020 (May to December) | 0 | 304 | 0 | 0 | 166 | 0 | 0 | 238 | 0 | 0 | 94 | 0 |

| | | | | | | | | | | | | |
|---|---|---|---|---|---|---|---|---|---|---|---|---|
| | (0.0) | (5.0) | (0.0) | (0.0) | (6.5) | (0.0) | (0.0) | (5.2) | (0.0) | (0.0) | (2.6) | (0.0) |
| 2021 (January to December) | 0<br>(0.0) | 4,456<br>(73.0) | 4,456<br>(100) | 0<br>(0.0) | 1,771<br>(69.8) | 1,771<br>(100) | 0<br>(0.0) | 3,438<br>(75.2) | 3,438<br>(100) | 0<br>(0.0) | 2,731<br>(76.4) | 2,731<br>(100) |
| 2022 (January to July) | 0<br>(0.0) | 832<br>(13.6) | 0<br>(0.0) | 0<br>(0.0) | 362<br>(14.3) | 0<br>(0.0) | 0<br>(0.0) | 638<br>(13.9) | 0<br>(0.0) | 0<br>(0.0) | 578<br>(16.2) | 0<br>(0.0) |
| **Comorbidity during baseline period, n (%)** | | | | | | | | | | | | |
| Down syndrome | 55<br>(0.4) | 15<br>(0.2) | 5<br>(0.1) | 41<br>(0.5) | 10<br>(0.4) | 3<br>(0.2) | 36<br>(0.4) | 9<br>(0.2) | 5<br>(0.1) | 38<br>(0.8) | 19<br>(0.5) | 16<br>(0.6) |
| Congenital heart disease | 404<br>(2.7) | 137<br>(2.2) | 87<br>(2.0) | 225<br>(2.7) | 61<br>(2.4) | 41<br>(2.3) | 115<br>(1.1) | 31<br>(0.7) | 25<br>(0.7) | 56<br>(1.2) | 31<br>(0.9) | 24<br>(0.9) |
| Bronchopulmonary dysplasia | 21<br>(0.1) | 1<br>(0.0) | 1<br>(0.0) | 15<br>(0.2) | 4<br>(0.2) | 2<br>(0.1) | 6<br>(0.1) | 2<br>(0.0) | 1<br>(0.0) | 4<br>(0.1) | 0<br>(0.0) | 0<br>(0.0) |
| Prematurity | 565<br>(3.8) | 177<br>(2.9) | 98<br>(2.2) | 243<br>(2.9) | 67<br>(2.6) | 49<br>(2.8) | 61<br>(0.6) | 18<br>(0.4) | 16<br>(0.5) | 11<br>(0.2) | 4<br>(0.1) | 3<br>(0.1) |
| Immunodeficiency | 20<br>(0.1) | 5<br>(0.1) | 4<br>(0.1) | 16<br>(0.2) | 3<br>(0.1) | 2<br>(0.1) | 14<br>(0.1) | 2<br>(0.0) | 2<br>(0.1) | 7<br>(0.2) | 4<br>(0.1) | 4<br>(0.1) |
| Pulmonary hypoplasia | 7<br>(0.0) | 0<br>(0.0) | 0<br>(0.0) | 1<br>(0.0) | 1<br>(0.0) | 0<br>(0.0) | 1<br>(0.0) | 0<br>(0.0) | 0<br>(0.0) | 0<br>(0.0) | 2<br>(0.1) | 2<br>(0.1) |
| Airway stenosis | 4<br>(0.0) | 3<br>(0.0) | 3<br>(0.1) | 3<br>(0.0) | 2<br>(0.1) | 2<br>(0.1) | 5<br>(0.0) | 4<br>(0.1) | 4<br>(0.1) | 2<br>(0.0) | 1<br>(0.0) | 1<br>(0.0) |
| Esophageal atresia | 1<br>(0.0) | 0<br>(0.0) | 0<br>(0.0) | 0<br>(0.0) | 0<br>(0.0) | 0<br>(0.0) | 2<br>(0.0) | 0<br>(0.0) | 0<br>(0.0) | 0<br>(0.0) | 0<br>(0.0) | 0<br>(0.0) |
| Congenital metabolic disorder | 39<br>(0.3) | 10<br>(0.2) | 5<br>(0.1) | 38<br>(0.5) | 19<br>(0.7) | 13<br>(0.7) | 39<br>(0.4) | 32<br>(0.7) | 22<br>(0.6) | 21<br>(0.5) | 27<br>(0.8) | 24<br>(0.9) |
| Neuromuscular disorder | 31<br>(0.2) | 9<br>(0.1) | 3<br>(0.1) | 26<br>(0.3) | 11<br>(0.4) | 3<br>(0.2) | 42<br>(0.4) | 17<br>(0.4) | 14<br>(0.4) | 35<br>(0.8) | 18<br>(0.5) | 13<br>(0.5) |
| Cystic fibrosis | 0 | 0 | 0 | 0 | 0 | 0 | 0 | 0 | 0 | 0 | 0 | 0 |

| | | | | | | | | | | | | |
|---|---|---|---|---|---|---|---|---|---|---|---|---|
| | (0.0) | (0.0) | (0.0) | (0.0) | (0.0) | (0.0) | (0.0) | (0.0) | (0.0) | (0.0) | (0.0) | (0.0) |
| No record of these comorbidities | 5,782 (94.7) | 4,263 (95.7) | 7,815 (94.0) | 2,391 (94.3) | 1,675 (94.6) | 9,869 (97.3) | 4,475 (97.8) | 3,361 (97.8) | 4,389 (96.7) | 3,479 (97.4) | 2,655 (97.2) | 5,782 (94.7) |
| **Palivizumab use, n (%)** | | | | | | | | | | | | |
| Yes | 343 (2.3) | 95 (1.6) | 47 (1.1) | 211 (2.5) | 59 (2.3) | 38 (2.1) | 174 (1.7) | 52 (1.1) | 37 (1.1) | 15 (0.3) | 13 (0.4) | 9 (0.3) |
| No record of use | 14,534 (97.7) | 6,011 (98.4) | 4,409 (98.9) | 8,100 (97.5) | 2,477 (97.7) | 1,733 (97.9) | 9,968 (98.3) | 4,522 (98.9) | 3,401 (98.9) | 4,522 (99.7) | 3,560 (99.6) | 2,722 (99.7) |
| **Size of healthcare facilities, n (%)** | | | | | | | | | | | | |
| Less than 200 beds | 679 (4.6) | 358 (5.9) | 255 (5.7) | 489 (5.9) | 196 (7.7) | 132 (7.5) | 524 (5.2) | 350 (7.7) | 254 (7.4) | 240 (5.3) | 302 (8.5) | 242 (8.9) |
| 200 to 499 beds | 8,137 (54.7) | 3,296 (54.0) | 2,395 (53.7) | 4,930 (59.3) | 1,501 (59.2) | 1,028 (58.0) | 5,994 (59.1) | 2,599 (56.8) | 1,962 (57.1) | 2,595 (57.2) | 1,982 (55.5) | 1,520 (55.7) |
| 500 or more beds | 6,061 (40.7) | 2,452 (40.2) | 1,806 (40.5) | 2,892 (34.8) | 839 (33.1) | 611 (34.5) | 3,624 (35.7) | 1,625 (35.5) | 1,222 (35.5) | 1,702 (37.5) | 1,289 (36.1) | 969 (35.5) |

*Rounding may create totals slightly over or below 100% in some cases.*

**Supplementary Table 8.** Baseline characteristics of children <5 years of age who are hospitalized with influenza in Japan by age group and COVID-19 era

| | Age <6 months | | | Age 6-11 months | | | Age 1-<2- years | | | Age 2-<5 years | | |
|---|---|---|---|---|---|---|---|---|---|---|---|---|
| | **Pre-COVID-19** | **During COVID-19** | **2021 season** | **Pre-COVID-19** | **During COVID-19** | **2021 season** | **Pre-COVID-19** | **During COVID-19** | **2021 season** | **Pre-COVID-19** | **During COVID-19** | **2021 season** |
| **Total number of patients per age group per time period, n (relative % by age group)** | 1,125 (20.4) | 155 (18.0) | 1 (16.7) | 568 (10.3) | 97 (11.3) | 1 (16.7) | 1,384 (25.1) | 275 (31.9) | 3 (50.0) | 2,431 (44.1) | 334 (38.8) | 1 (16.7) |
| **Sex, n (%)** | | | | | | | | | | | | |
| Female | 528 (46.9) | 63 (40.6) | 1 (100) | 260 (45.8) | 38 (39.2) | 1 (100) | 620 (44.8) | 118 (42.9) | 0 (0.0) | 1,035 (42.6) | 126 (37.7) | 0 (0.0) |
| Male | 597 (53.1) | 92 (59.4) | 0 (0.0) | 308 (54.2) | 59 (60.8) | 0 (0.0) | 764 (55.2) | 157 (57.1) | 3 (100) | 1,396 (57.4) | 208 (62.3) | 1 (100) |
| **Age, months** | | | | | | | | | | | | |
| **Mean (SD)** | 2.6 (1.2) | 2.6 (1.3) | 0.0 (.) | 8.6 (1.7) | 8.5 (1.7) | 10.0 (.) | 17.3 (3.4) | 17.3 (3.3) | 19.0 (1.7) | 40.2 (10.6) | 40.3 (10.6) | 27.0 (.) |
| **Median (IQR)** | 2.0 (2.0-3.0) | 3.0 (2.0-4.0) | 0.0 (0.0-0.0) | 9.0 (7.0-10.0) | 9.0 (7.0-10.0) | 10.0 (10.0-10.0) | 17.0 (14.0-20.0) | 17.0 (14.0-20.0) | 20.0 (17.0-20.0) | 40.0 (31.0-49.0) | 40.0 (31.0-49.0) | 27.0 (27.0-27.0) |
| **Season by year, n (%)** | | | | | | | | | | | | |
| 2016 (May 2016 to April 2017) | 255 (22.7) | 0 (0.0) | 0 (0.0) | 108 (19.0) | 0 (0.0) | 0 (0.0) | 276 (19.9) | 0 (0.0) | 0 (0.0) | 485 (20.0) | 0 (0.0) | 0 (0.0) |
| 2017 (May 2017 to April 2018) | 303 (26.9) | 0 (0.0) | 0 (0.0) | 174 (30.6) | 0 (0.0) | 0 (0.0) | 396 (28.6) | 0 (0.0) | 0 (0.0) | 804 (33.1) | 0 (0.0) | 0 (0.0) |
| 2018 (May 2018 to April 2019) | 423 (37.6) | 0 (0.0) | 0 (0.0) | 186 (32.7) | 0 (0.0) | 0 (0.0) | 490 (35.4) | 0 (0.0) | 0 (0.0) | 796 (32.7) | 0 (0.0) | 0 (0.0) |
| 2019 (May 2019 to April 2020) | 144 (12.8) | 152 (98.1) | 0 (0.0) | 100 (17.6) | 94 (96.9) | 0 (0.0) | 222 (16.0) | 269 (97.8) | 0 (0.0) | 346 (14.2) | 333 (99.7) | 0 (0.0) |

| | | | | | | | | | | | | |
|---|---|---|---|---|---|---|---|---|---|---|---|---|
| 2020 (May to December) | 0 (0.0) | 1 (0.6) | 0 (0.0) | 0 (0.0) | 2 (2.1) | 0 (0.0) | 0 (0.0) | 0 (0.0) | 0 (0.0) | 0 (0.0) | 0 (0.0) | 0 (0.0) |
| 2021 (January to December) | 0 (0.0) | 1 (0.6) | 1 (100) | 0 (0.0) | 1 (1.0) | 1 (100) | 0 (0.0) | 3 (1.1) | 3 (100) | 0 (0.0) | 1 (0.3) | 1 (100) |
| 2022 (January to July) | 0 (0.0) | 1 (0.6) | 0 (0.0) | 0 (0.0) | 0 (0.0) | 0 (0.0) | 0 (0.0) | 3 (1.1) | 0 (0.0) | 0 (0.0) | 0 (0.0) | 0 (0.0) |
| **Comorbidity during baseline period, n (%)** | | | | | | | | | | | | |
| Down syndrome | 1 (0.1) | 1 (0.6) | 0 (0.0) | 1 (0.2) | 1 (1.0) | 0 (0.0) | 0 (0.0) | 0 (0.0) | 0 (0.0) | 4 (0.2) | 0 (0.0) | 0 (0.0) |
| Congenital heart disease | 23 (2.0) | 1 (0.6) | 0 (0.0) | 16 (2.8) | 4 (4.1) | 0 (0.0) | 13 (0.9) | 2 (0.7) | 0 (0.0) | 8 (0.3) | 1 (0.3) | 0 (0.0) |
| Bronchopulmonary dysplasia | 1 (0.1) | 0 (0.0) | 0 (0.0) | 1 (0.2) | 0 (0.0) | 0 (0.0) | 1 (0.1) | 0 (0.0) | 0 (0.0) | 0 (0.0) | 0 (0.0) | 0 (0.0) |
| Prematurity | 19 (1.7) | 3 (1.9) | 0 (0.0) | 16 (2.8) | 6 (6.2) | 0 (0.0) | 1 (0.1) | 1 (0.4) | 0 (0.0) | 1 (0.0) | 0 (0.0) | 0 (0.0) |
| Immunodeficiency | 1 (0.1) | 0 (0.0) | 0 (0.0) | 4 (0.7) | 1 (1.0) | 0 (0.0) | 3 (0.2) | 1 (0.4) | 0 (0.0) | 8 (0.3) | 0 (0.0) | 0 (0.0) |
| Pulmonary hypoplasia | 0 (0.0) | 1 (0.6) | 0 (0.0) | 0 (0.0) | 0 (0.0) | 0 (0.0) | 0 (0.0) | 0 (0.0) | 0 (0.0) | 0 (0.0) | 0 (0.0) | 0 (0.0) |
| Airway stenosis | 0 (0.0) | 0 (0.0) | 0 (0.0) | 0 (0.0) | 0 (0.0) | 0 (0.0) | 1 (0.1) | 1 (0.4) | 0 (0.0) | 0 (0.0) | 0 (0.0) | 0 (0.0) |
| Esophageal atresia | 0 (0.0) | 0 (0.0) | 0 (0.0) | 0 (0.0) | 0 (0.0) | 0 (0.0) | 0 (0.0) | 0 (0.0) | 0 (0.0) | 0 (0.0) | 0 (0.0) | 0 (0.0) |
| Congenital metabolic disorder | 1 (0.1) | 0 (0.0) | 0 (0.0) | 4 (0.7) | 0 (0.0) | 0 (0.0) | 11 (0.8) | 2 (0.7) | 0 (0.0) | 25 (1.0) | 0 (0.0) | 0 (0.0) |
| Neuromuscular disorder | 0 (0.0) | 0 (0.0) | 0 (0.0) | 5 (0.9) | 3 (3.1) | 0 (0.0) | 17 (1.2) | 2 (0.7) | 0 (0.0) | 28 (1.2) | 1 (0.3) | 0 (0.0) |

| | | | | | | | | | | | | |
|---|---|---|---|---|---|---|---|---|---|---|---|---|
| Cystic fibrosis | 0 (0.0) | 0 (0.0) | 0 (0.0) | 0 (0.0) | 0 (0.0) | 0 (0.0) | 0 (0.0) | 0 (0.0) | 0 (0.0) | 0 (0.0) | 0 (0.0) | 0 (0.0) |
| No record of these comorbidities | 1,082 (96.2) | 150 (96.8) | 1 (100.0) | 529 (93.1) | 85 (87.6) | 1 (100.0) | 1,339 (96.7) | 269 (97.8) | 3 (100.0) | 2,367 (97.4) | 332 (99.4) | 1 (100.0) |
| **Palivizumab use, n (%)** | | | | | | | | | | | | |
| Yes | 11 (1.0) | 2 (1.3) | 0 (0.0) | 15 (2.6) | 5 (5.2) | 0 (0.0) | 17 (1.2) | 1 (0.4) | 0 (0.0) | 2 (0.1) | 1 (0.3) | 0 (0.0) |
| No record of use | 1,114 (99.0) | 153 (98.7) | 1 (100) | 553 (97.4) | 92 (94.8) | 1 (100) | 1,367 (98.8) | 274 (99.6) | 3 (100) | 2,429 (99.9) | 333 (99.7) | 1 (100) |
| **Size of healthcare institution, n (%)** | | | | | | | | | | | | |
| Less than 200 beds | 35 (3.1) | 2 (1.3) | 0 (0.0) | 19 (3.3) | 1 (1.0) | 0 (0.0) | 71 (5.1) | 14 (5.1) | 0 (0.0) | 93 (3.8) | 13 (3.9) | 0 (0.0) |
| 200 to 499 beds | 595 (52.9) | 76 (49.0) | 1 (100) | 299 (52.6) | 52 (53.6) | 1 (100) | 776 (56.1) | 143 (52.0) | 2 (66.7) | 1,330 (54.7) | 175 (52.4) | 1 (100) |
| 500 or more beds | 495 (44.0) | 77 (49.7) | 0 (0.0) | 250 (44.0.) | 44 (45.4) | 0 (0.0) | 537 (38.8) | 118 (42.9) | 1 (33.3) | 1,008 (41.5) | 146 (43.7) | 0 (0.0) |

*Rounding may create totals slightly over or below 100% in some cases.*
